# Integrative Brain Transcriptome Analysis Links Complement Component 4 and *HSPA2* to the *APOE* ε2 Protective Effect in Alzheimer Disease

**DOI:** 10.1101/2020.11.23.20235762

**Authors:** Rebecca Panitch, Junming Hu, Jaeyoon Chung, Congcong Zhu, Gaoyuan Meng, Weiming Xia, David A. Bennett, Kathryn L. Lunetta, Tsuneya Ikezu, Rhoda Au, Thor D. Stein, Lindsay A. Farrer, Gyungah R. Jun

**Author notes:** Address Correspondence to: Lindsay A. Farrer, PhD, Department of Medicine (Biomedical Genetics), Boston University School of Medicine, 72 East Concord Street, Boston, MA 02118, USA 02118,; Gyungah R. Jun, PhD, Department of Medicine (Biomedical Genetics), Boston University School of Medicine, 72 East Concord Street, Boston, MA 02118, USA 02118,.

## Abstract

Mechanisms underlying the protective effect of apolipoprotein E (*APOE*) ε2 against Alzheimer’s disease (AD) are not well understood. We analyzed gene expression data derived from autopsied brains donated by 982 individuals including 135 *APOE* ε 2/ε 3 carriers. Complement pathway genes *C4A* and *C4B* were among the most significantly differentially expressed genes between ε 2/ε 3 AD cases and controls. We also identified an *APOE* ε2/ε3 AD-specific co-expression network enriched for astrocytes, oligodendrocytes and oligodendrocyte progenitor cells containing the genes *C4A, C4B*, and *HSPA2*. These genes were significantly associated with the ratio of phosphorylated tau at position 231 to total Tau but not with amyloid-β 42 level, suggesting this *APOE* ε 2 related co-expression network may primarily be involved with tau pathology. *HSPA2* expression was oligodendrocyte specific and significantly associated with C4B protein. Our findings provide the first evidence of a crucial role of the complement pathway in the protective effect of *APOE* ε2 for AD.

## Introduction

Apolipoprotein E (*APOE*) genotype, the strongest genetic risk factor for late onset Alzheimer disease (AD), is determined by the combination of amino acids at positions at 112 and 158 yielding three common alleles (ε2, ε3, and ε4) [1,2]. Among persons of European ancestry, a single copy of the ε4 allele is associated with a 3-4 fold increased risk of AD and ε4 homozygotes have a 10-12 fold increased risk compared to those with the common ε3/ε3 genotype [3,4,5]. In persons of European ancestry, the *APOE* ε2 allele is associated with a 60% and 90% decreased risk of AD among heterozygotes and homozygotes, respectively, compared to the ε3/ε3 genotype [3,6]. By comparison, the AD/ε4 association is greater among East Asians and attenuated in African Americans [3,5]. Population differences in the frequency of ε4 may account for some of the ε4 effect variability, but other factors are likely involved including the modifying effect of a polymorphic variant in the *APOE* promoter region which has been shown to influence *APOE* expression *in vitro* [5].

The opposing effects of ε2 and ε4 on AD risk have been related to similar mechanisms such as amyloid-β (Aβ) aggregation and clearance and neurofibrillary tangle formation [7,8], however, studies are contradictory or ambiguous about the relationship of these effects, especially the less studied ε2 protective effect, with AD-related pathology [9]. In brains with an autopsy-confirmed diagnosis of AD without evidence of other neurodegenerative disease, ε2 is generally associated with decreased neurofibrillary tangles and with reduced neuritic plaques [10], hallmarks of AD neuropathology [11], except in elderly subjects (aged 90 or older) where ε2 is associated with increased neuritic plaques [12]. The mechanism underlying the ε4-associated risk has been linked to many AD pathways including Aβ aggregation and lipid metabolism, and AD brains with the *APOE* ε4 allele and without other pathologies are associated with increased neuritic plaques but not associated with neurofibrillary tangles [2,10,13,14]. While the ε4 AD risk mechanism has been extensively researched, the biological underpinning of the protective effect of ε2 is not well understood.

The advent of next-generation sequencing technology has enabled transcriptomic studies (e.g., differential expression analyses, alternative splicing, and gene expression networks [15,16]) using RNA-sequencing (RNA-seq) data derived from neuropathologically evaluated brain tissue [17]. Multiple studies have linked expression of AD-associated genes to several pathways in the brain including synapse function, cytoskeleton structure, and immune function [18,19]. We analyzed differential gene expression, gene expression networks, and immunoassay levels of AD-related proteins in brain tissue from pathologically confirmed AD cases and controls according to *APOE* genotype to identify genes and biological pathways that may be functionally involved in the mechanism underlying the protective effect of *APOE* ε2.

## Materials and Methods

### Sources of Brain Transcriptomic and Phenotypic Data

Publicly available RNA sequencing and neuropathological data were obtained from the CommonMind Consortium portal (http://www.synapse.org) including preprocessed, quality controlled, and normalized gene expression data derived from dorsolateral prefrontal cortex area tissue donated by 627 participants (380 autopsy-confirmed AD cases and 247 controls) of the Religious Orders Study and Rush Memory and Aging Project (ROSMAP; https://www.radc.rush.edu) [20] and from temporal cortex area tissue donated by 162 participants (82 autopsy-confirmed AD cases and 80 controls) of the Mayo Clinic Study of Aging (MAYO) [21,22,23]. AD diagnosis was established according to the National Institute of Aging (NIA)

Reagan criteria for intermediate or high probability of AD[24]. Available neuropathological measures for ROSMAP participants included Braak staging for neurofibrillary tangles and the Consortium to Establish a Registry for Alzheimer Disease (CERAD) semi-quantitative criteria for neuritic plaques (CERAD Score) [25]. No neuropathological data were available for MAYO participants. *APOE* genotype information was available for all subjects. Gene expression levels were quantified as normalized fragments per kilobase of transcript per million (FPKM) reads in the ROSMAP dataset and as normalized gene counts in the MAYO dataset.

We obtained frontal cortex tissue specimens from 208 participants (64 autopsy-confirmed AD cases and 129 controls) of the Framingham Heart Study (FHS) and Boston University Alzheimer’s Disease Center (BUADC) that were examined by the Neuropathology Core of the BUADC. The FHS is a community-based multi-generational longitudinal study of health that surveilles participants for cognitive decline and dementia using protocols described elsewhere [26]. Brain tissue was collected after death from 184 FHS participants who enrolled in the brain donation with informed consent from the next of kin. The second cohort consisted of 24 participants from the BUADC with and without cognitive impairment who prior to death underwent annual cognitive evaluations using the National Alzheimer’s Disease Coordinating Center (NACC) Uniform Data Set (UDS) protocol [27]. Neuropathological assessment was performed following procedures and criteria established by the Department of Veterans Affairs-Boston University Brain bank [28]. AD was diagnosed using the NIA Reagan criteria. Braak stages were assigned using the same criteria as described above and CERAD scores were derived using semi-quantitative criteria for neuritic plaques [29]. Neuritic plaques were defined as plaques with argyrophilic dystrophic neurites, with or without dense amyloid cores. This study was approved by the institutional review boards from Boston University Medical Center and the Edith Nourse Rogers Memorial Veterans Hospital, Bedford, MA.

### RNA Library Preparation, Sequencing, and Sample QC

Total RNAs from the dorsolateral prefrontal cortex (Brodmann area 8/9) of 208 brains from the FHS/BUADC study were extracted using the Promega Maxwell RSC simplyRNA Tissue Kit (Cat No# AS1340) according to the manufacturer’s protocol. The integrity and quality of RNA (RNA integrity number, RIN) was determined using the High Sensitivity RNA Screen Tape Assay run on an Agilent 2200 Tape Station (Agilent Technologies, Palo Alto, CA). After excluding brain samples with RIN < 5, brain samples were randomized into seven library batches based on diagnosis, *APOE* genotype, sex, and RIN. Since there were only seven samples from AD cases with *APOE* genotypes 2/2 or 2/3, these specimens were included in batches 1 to 3 only. The BU Microarray & Sequencing Resource Core performed RNA sequencing (RNA-seq) library preparation. The libraries were prepared from total RNA enriched for mRNA using NEBNext Poly(A) mRNA Magnetic Isolation Module and NEBNext Ultra II Directional RNA Library Preparation Kit for Illumina (New England Biolabs, USA) and sequenced on an Illumina NextSeq 500 instrument (Illumina, USA). Only 193 of the 208 samples containing AD status were used in downstream analysis.

### Mapping, Quality Control, and Quantification of Gene Expression

RNA-seq data from 193 FHS/BUADC brains were processed using an automated pipeline. Quality controls (QC) of the sequence data including checks for over-abundance of adaptors and over-represented sequence was performed using FastQC [30]. Low-quality reads (5% of the total) were filtered out using the *Trimmomatic* option (version 0.39), which is a fast, multithreaded command line tool to trim and crop Illumina (FASTQ) data and remove adapters [31]. After trimming adapter sequences, reads passing initial QC were aligned to the human reference genome (GRCh38.95) using STAR (version 2.6.1c) which implements 2-pass mapping to increase mapping chances of splice reads from novel junctions [32,33]. We used the *readFilesCommand* option for reading input files, the *TranscriptomeSAM* option for mapping reads translated into transcript coordinates, and the *GeneCounts* option for counting mapped reads per gene under the mapping mode set to *quantMode* and then *twoPassMode* options. This process produced a BAM file of mapped paired-end reads for each sample with a corresponding alignment report file.

Post alignment quality of BAM files was evaluated for gene coverage and junction saturation using the RseQC (version 3.0.0, python3) [34]. The RSeQC program evaluates uniformity of coverage over entire genes using the *gene_Body_coverage* option by checking if inner distance between read pairs is within expecting range of fragments lengths using the *inner_distance* option, and ensuring sequencing depth using the *junction_saturation* option. Gene and isoform levels were quantified using RSEM (version 1.3.1) and Bowtie2 (version 2.3.4.1) and annotated using Homo_sapiens. GRCh38.95.gtf annotation files. This process generated gene or isoform expression data for each sample containing gene id, gene length, effective gene length, expected count, counts per million (CPM), and fragments per kilobase of exon model per million reads mapped (FPKM) reads. Batch effects of seven different library and three sequencing batches with the quantified gene expression data were investigated using principle component analysis (PCA). We did not detect significant batch effects for both library and sequencing batches (**Supplementary Fig. 1**).

### Differential Gene Expression Analysis

Evaluation of differential gene expression between AD and control brains was performed using the LIMMA software, which can accommodate pre-normalized expression values [35]. After excluding genes with no FPKM reads or gene counts in the MAYO and ROSMAP datasets and genes with FPKM reads < 5 in the FHS/BUADC dataset, expression of each gene/isoform was compared between AD cases and controls using linear regression models including the log-transformed normalized expression values and terms for age of death (AOD), sex, and RIN. The number of genes remaining after filtering was 42,413 in the ROSMAP dataset, 47,205 in the MAYO dataset, and 33,476 in the FHS/BUADC dataset. Models analyzing FHS/BUADC data also included batch number, and models for ROSMAP data included batch number, education level, and post-mortem interval (PMI). The ROSMAP sample was reduced to 568 specimens (339 AD cases and 229 controls) due to missing RIN. Analyses were conducted in the total sample and separately in *APOE* genotype groups (ε2/3, ε3/3, and ε3/4). There were insufficient numbers of ε2/2 or ε4/4 subjects for analyses within these genotype groups. Results across datasets and *APOE* genotype groups within datasets were combined by a sample size-weighted meta-analysis with log2 of fold change (logFC) as direction using the software METAL [36]. Only genes included in all three datasets were considered for follow-up analysis. Significance thresholds were set according to the number of genes tested in each *APOE* genotype group and in the total sample: ε2/ε3 (p=2.35×10^−6^), ε3/ε3 (p=1.96×10^−6^), ε3/ε4 (p=2.42×10^−6^), and total (p=1.82×10^−6^).

### Association Analysis of Gene Expression Levels on Neuropathological Traits

The distribution of Braak Stage and CERAD Score in the ROSMAP and FHS/BUADC datasets by *APOE* genotype is shown in Supplemental Table 6. Values for each trait were adjusted for age at death and sex, and the residuals were rank-transformed as previously described [37]. Association of log2-transformed expression levels for the top-ranked differentially expressed genes (DEGs) with each rank-transformed neuropathological trait was evaluated in the total sample using linear regression models that adjusted for PMI and RIN in the ROSMAP dataset and for RIN in the FHS/BUADC dataset. Only the 568 ROSMAP samples including RIN were included in this analysis. We also ran the same linear regression models in the *APOE* subgroups (ε2/3, ε3/3, and ε3/4) exclusively in ROSMAP due to insufficient sample sizes in FHS/BUADC (**Supplementary Table 6**). Results obtained for the two datasets analyzed separately were combined by meta-analysis using the inverse variance model.

### Gene Co-Expression Network and Enrichment Analyses

Gene co-expression analysis was performed using the weighted correlation network analysis (WGCNA) in R package [38]. We created a signed adjacency matrix with a soft-power parameter determined by reaching a scale-free topological fit to at least 0.8 and maximizing mean connectivity. Soft-power creates a fuzzy threshold for gene connectivity which reduces network connectivity noise. We performed hierarchical clustering using a dissimilatory topological overlap matrix (TOM) which was created based on the adjacency matrix. Labels were then assigned to networks using the Dynamic tree cut package with a minimal network size of 100 genes [39]. Networks with highly correlated eigenvalues were merged using the mergeCloseModules function with a height of 0 [38]. The signedKME function was used to assign fuzzy membership values to all genes in each network. This network-building pipeline was applied to each dataset in six sample partitions defined by *APOE* genotype and AD status. The soft-power parameter values in these partitions were determined by maximizing scale-free topological fit and median connectivity. The soft-power parameter values for FHS/BUADC partitions were assigned as ε23 controls=6, ε23 AD cases=16, ε33 controls=12, ε33 AD cases=7, ε34 controls=12, and ε34 AD cases=8. Soft-power parameter values for all partitions in the ROSMAP and the MAYO datasets were set as 12. Network preservation across datasets with the same *APOE* genotype and AD status was evaluated using Z_Summary_ statistics which were calculated using the modulePreservation function in the WGCNA package [40]. Z_Summary_ values > 10 were considered as highly preserved networks between two datasets.

Gene enrichment analysis was conducted for network networks derived from subsets of the total sample stratified by *APOE* genotype and AD status using the userListEnrichment function in WGCNA. Networks from the ROSMAP dataset that were highly preserved in at least one other dataset were tested for AD gene enrichment using the gene-lists of DEGs from the *APOE* subgroup and total sample analyses as well as previously known AD genes from a recent genome-wide association study [1]. These AD gene-lists included genes from the differential expression analysis (p<0.01) corresponding to the network’s *APOE* genotype group that were also moderately differentially expressed in the total sample (p<0.01), genes that were differentially expressed between AD cases and controls in the total sample (p<10^−6^), and genes that showed modest evidence for association with AD (p<10^−3^) in a recent large GWAS [1]. P-values were adjusted using the Bonferroni method to correct for the number of separate analyses.

### Brain Cell-Type Specific Expression Profiles and Enrichment Analysis

Raw FASTQ single-nucleus RNA sequencing data derived from prefrontal cortex (Brodmann area 10) of 48 subjects (24 AD cases and 24 controls) in the ROSMAP dataset were obtained from the Synapse database. Read counts were aligned to a reference genome (GRCh38) by CellRanger software (v.3.1.0). We used a cut-off value of 200 for unique molecular identifiers (UMI) for better detection of the microglia populations due to their small representation in the dataset [41]. Genes without unique names, with low expression across all cell types, and that do not encode proteins were excluded. We included all AD genes contained in the co-expression networks derived from analyses described above regardless of single-nuclei expression levels, including genes from AD GWAS annotated to associated SNPs (p<10^−3^) [1], DEGs identified in the corresponding to *APOE* genotype group analysis (p<0.01) that were also DEGs in the total sample (p<0.01), and DEGs identified in the total sample (p<10^−6^). Data including 69,918 nuclei with 5,578 genes after filtering were normalized and clustered using Seurat (v.3.0.0).

Data were further processed using the NormalizeData, FindVariableFeatures, and ScaleData functions. Gene expression measures were scaled by 10,000 multiplied by the total library size and then log-transformed. The ScaleData method was used to adjust for the total number of counts. We conducted principle component (PC) analysis to reduce the model complexity of the high variable expression of 3,179 genes (marker genes). A cell type-specific expression metric was calculated by dividing the expression in each cell type by the total expression across all cell types. The top 10 PCs were included in t-SNE analysis. For each cluster, cell-type labels were assigned by known marker genes [41]. This procedure yielded eight cell type-specific clusters representing astrocytes, endothelial cells, excitatory neurons, inhibitory neurons, microglia, oligodendrocytes, oligodendrocyte progenitor cells, and pericytes. Endothelial cells and pericytes were not included in subsequent analysis due to their low proportion in the dataset. The average expression for each cell type per gene was calculated using the AverageExpression package in Seurat. This function calculates the average expression for the exponential of raw counts minus 1. Log transformed fold changes (Log2FC) in expression between AD and control samples were evaluated for enriched genes in *APOE* ε2/ε3 AD network within each cell type using the Wilcoxon rank-sum test. False discovery rate (FDR) p-values were adjusted using the Benjamini-Hochberg procedure implemented in R [42]. Novel marker genes were determined for each cell-type cluster using the FindAllMarkers function in Seurat with default parameters. Marker genes that were differentially expressed in a given cell-type with a false discovery rate (FDR) adjusted p-value > 0.05 were excluded from subsequent analyses. Cell-type enrichment analysis was conducted for each network using the userListEnrichment function in WGCNA and cluster markers identified for each cell-type. All enrichment p-values were corrected for the number of separate analyses using the Bonferroni method.

### Immunoassay Measurement and Their Relationship with Gene Expression

Frozen tissue from the dorsolateral prefrontal cortex (Brodmann area 8/9) of the 208 FHS/BUADC autopsied brains was weighed and placed on dry ice. Freshly prepared, ice cold 5M Guanidine Hydrochloride in Tris-buffered saline (20 mM Tris-HCl, 150 mM NaCl, pH 7.4) containing 1:100 Halt protease inhibitor cocktail (Thermo Fischer Scientific, Waltham, MA) and 1:100 Phosphatase inhibitor cocktail 2 & 3 (Sigma-Aldrich, St. Louis, MO) was added to the brain tissue at 5:1 (5M Guanidine Hydrochloride volume (ml):brain wet weight (g)) and homogenized with Qiagen Tissue Lyser LT at 50Hz for five min. The homogenate was then mixed (regular rocker) overnight at room temperature. The lysate was diluted with 1% Blocker A (Meso Scale Discovery (MSD), Rockville, Maryland, #R93BA-4) in wash buffer according to specific immunoassays: 1:300 for total tau, pTau231 (MSD #K15121D-2), pTau181, and PSD-95 (MSD #K250QND), and 1:4000 for beta-amyloid 1-42 (MSD #K15200E-2). Samples were subsequently centrifuged at 17,000g and 4°C for 15 minutes, after which the supernatant was applied to the immunoassays. To capture tau phosphorylated at Thr residue 181, antibody AT270 was used and the detecting antibody was the biotinylated HT7 that recognizes residue 159–163 of tau (Thermo Scientific, Rockford, IL). Internal calibrators of p-tau and tau were used (MSD) [43]. Standards with known concentrations were used for Aβ. For PSD-95, arbitrary values were assigned to a reference brain lysate, which was run as a standard curve with every plate. All standards and samples were run in duplicate. Measurements were performed using the multi-detection SPECTOR 6000 Imager (MSD).

For immunoassays of C4A and C4B, ice cold PBS buffer (Gibco, ref#10010-023) was added to the brain tissue at 5:1 (PBS in ml vs brain wet weight in gram), and homogenized with Qiagen Tissue Lyser LT at 50Hz for five min. The homogenate was centrifuged at 17,000g and 4°C for 15 minutes, then the supernatant was aliquoted and stored at -80°C. Lysate was diluted 1:200 (C4A) or left undiluted (C4B) and applied to ELISA assays according to the manufacturer’s protocol (C4A/C4B: Novus Biologicals, NBP2-70043 & NBP2-70046). Absorbance was measured at 450 nm using Biotek Synergy HT microplate reader.

We evaluated association of DEGs with rank-transformed immunoassay measures of Aβ42, pTau181/tTau ratio, pTau231/tTau ratio, C4A, and C4B that were adjusted for age at death and sex. We included only the 193 brain samples with information on AD case status in our downstream analysis. Analyses included DEGs significant in the ε2/ε3 group (*C4A, C4B, GFAP, NPNT)*, or from the M01 gene network and significant DEGs in oligodendrocytes (*HSPA2, PHLPP1, DOCK1*, and *LPAR1*). Gene expression values were log2-transformed and tested for association with the level of each protein using linear regression models adjusted for an additional covariate, RIN.

## Results

### Differentially Expressed Genes in *APOE* Subgroups

Gene expression was examined in post-mortem tissue from the prefrontal cortex area of 627 participants in the Religious Order Study or the Rush Memory and Aging Project (ROSMAP) and 193 participants in the Framingham Heart Study or Boston University Alzheimer’s Disease Center (FHS/BUADC), and from the temporal cortex area of 162 participants in the Mayo Clinic Study of Aging (MAYO) (**Supplementary Table 1 and Supplementary Figure 1**). The combined sample had the following distribution of *APOE* genotypes: ε2/ε3 with 48 AD cases and 87 controls, ε3/ε3 with 280 AD and 302 controls, and ε3/ε4 with 169 AD cases and 45 controls (**Supplementary Table 1**). We identified 1,114 genes with differences in expression between AD cases and controls in the total sample that were transcriptome-wide significant (p<10^−6^) (**Supplementary Table 2 and Supplementary Figures 2-4**). All of the top 20 differentially expressed genes (DEGs) in the total sample were up-regulated in AD compared to controls including *EMP3* (p=1.6×10^−18^), *NPNT* (p=9.3×10^−18^), and *SLC4A11* (p=1.2×10^−17^). The genes showing the largest differences in expression between AD cases and controls varied by *APOE* genotypes. Among the transcriptome-wide significant results in the total sample, the most significant expression differences in the *APOE* ε2/ε3 subgroup (p<10^−3^) were observed with *C4A, C4B, GFAP* and *NPNT* each of which was up-regulated in AD cases compared to controls (**Table 1 and Fig. 1A**). These genes were consistently up-regulated in each dataset (**Supplementary Table 3**). Expression differences of *C4A* and *C4B* were progressively weaker in the ε3/ε3 and ε3/ε4 groups, respectively (**Table 1 and Fig. 1A**). *GFAP* and *NPNT* showed at least nominally significant differences in expression among ε3/ε3 but not ε3/ε4 individuals (**Table 1**).

**Table 1.** **Genes Differentially Expressed Between AD Cases and Controls Among *APOE ε2/ε3* Subjects**

| Gene * | *APOE* $\epsilon 2/\epsilon 3$ | | *APOE* $\epsilon 3/\epsilon 3$ | | *APOE* $\epsilon 3/\epsilon 4$ | | Total | |
| --- | --- | --- | --- | --- | --- | --- | --- | --- |
|  | Z | P-value | Z | P-value | Z | P-value | Z | P-value |
| *C4A* | 4.28 | $1.9 \times 10^{-5}$ | 2.78 | $5.4 \times 10^{-3}$ | 2.48 | 0.01 | 5.73 | $1.0 \times 10^{-8}$ |
| *C4B* | 4.10 | $4.2 \times 10^{-5}$ | 3.50 | $4.7 \times 10^{-4}$ | 2.17 | 0.03 | 6.08 | $1.2 \times 10^{-9}$ |
| *GFAP* | 4.12 | $3.8 \times 10^{-5}$ | 4.03 | $5.6 \times 10^{-5}$ | 1.51 | 0.13 | 6.89 | $5.4 \times 10^{-12}$ |
| *NPNT* | 3.65 | $2.6 \times 10^{-4}$ | 6.34 | $2.4 \times 10^{-10}$ | 1.58 | 0.11 | 8.58 | $9.3 \times 10^{-18}$ |

\* Surpassed significance thresholds of $p < 10^{-3}$ in the *APOE* $2\epsilon/\epsilon 3$ group and $p < 10^{-6}$ in the total sample.

**Fig. 1.**
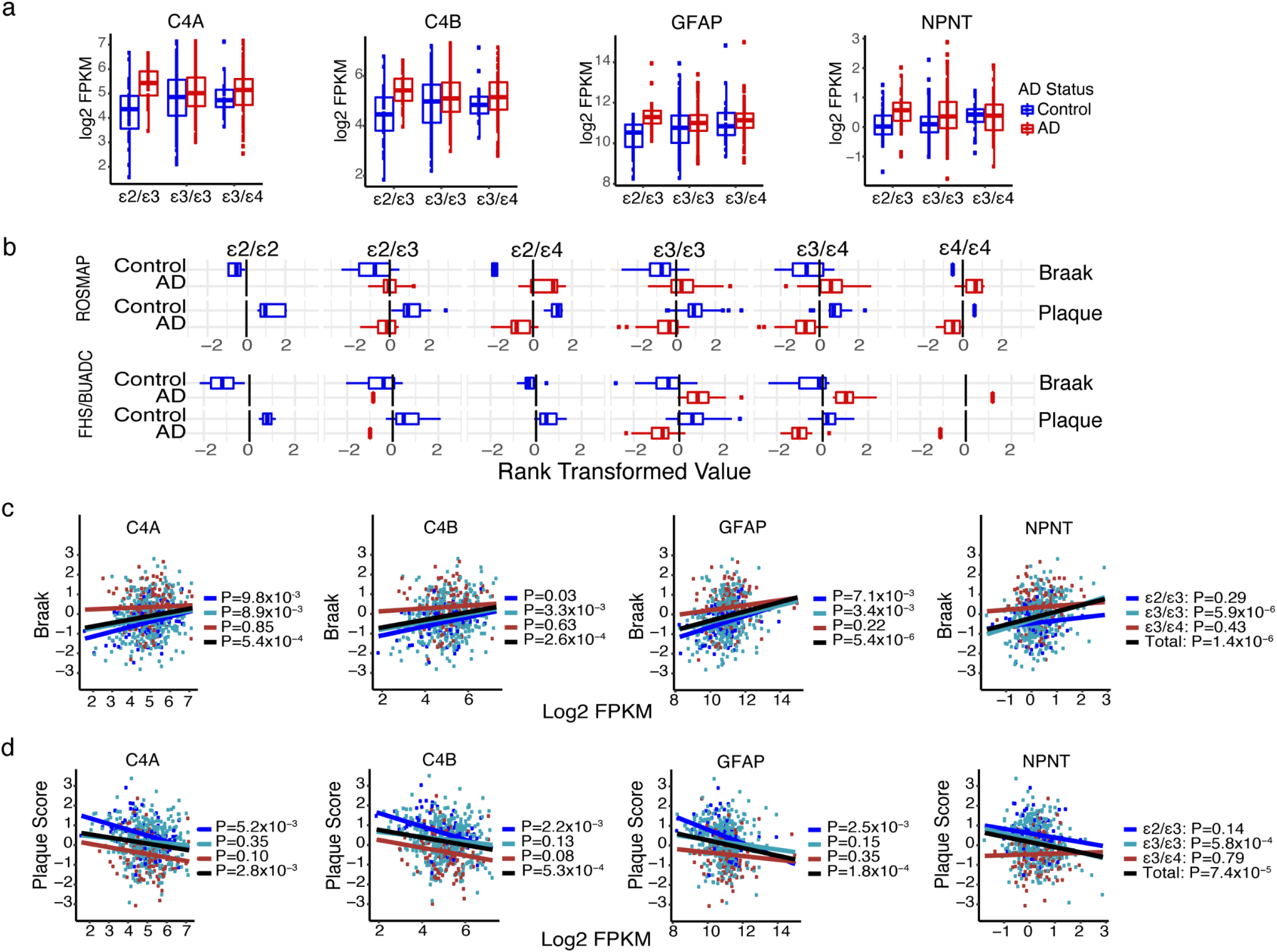
Differentially expressed genes (DEGs) among *APOE* ε2/ε3 individuals and in the total sample. **(a)** Boxplots showing distribution of gene expression level (represented as log2 FPKM) by AD status and *APOE* genotype for top-ranked DEGs among *APOE* ε2/ε3 subjects in the ROSMAP dataset. **(b)** Boxplots showing the distribution of rank-transformed plaque score and Braak stage by *APOE* genotype and AD status among subjects in the ROSMAP and FHS/BUADC datasets. **(c and d)** Scatterplots showing the correlation of expression of the top-ranked DEGs among *APOE* ε2/ε3 subjects with rank-transformed Braak stage **(c)** and plaque score **(d)** in the ROSMAP dataset according to *APOE* genotype depicted by color (ε2/ε3=dark blue, ε3/ε3=light blue, ε3/ε4=red, total=black). Coordinates for expression (quantified as log2 FPKM) plotted against plaque or Braak score for each subject are shown as dots and their correlation across subjects is represented by fitted solid lines.

Eleven of the 1,114 transcriptome-wide significant DEGs in the total sample were also moderately significant (p<0.01) in each *APOE* genotype subgroup (**Supplementary Figure 5 and Supplementary Table 4**). The most significant gene in this group is *C1QTNF5* which encodes complement C1q tumor necrosis factor-related protein 5 (p=2.8×10^−17^). Contribution of these genes to AD may be independent of *APOE* genotype.

### Expression of Top DEGs in *APOE* ε2/ε3 Subjects is Associated With AD-Related Neuropathology

Primary neuropathological hallmarks of AD in the ROSMAP and FHS/BUADC datasets, density of neurofibrillary tangles (NFT) measured by Braak Stage and neuritic plaques measured according to the Consortium to Establish a Registry for Alzheimer Disease (CERAD) Score, were comparable across the two datasets (**Fig. 1B** and **Supplementary Table 5**). Expression of all four top-ranked DEGs in the *APOE* ε2/ε3 subgroup (*C4A, C4B, GFAP*, and *NPNT*) was moderately associated (P<0.01) with both Braak stage and plaque score in the combined datasets (**Fig. 1C and Supplementary Table 6**). Expression of *C4A, C4B*, and *GFAP* in the ROSMAP dataset was at least nominally associated with Braak stage among ε2/ε3 and ε3/ε3 subjects and with neuritic plaque density in ε2/ε3 subjects only, whereas expression of *NPNT* was significantly associated with these traits only among ε3/ε3 subjects (**Fig. 1C, Fig. 1D, and Supplementary Table 7**).

### AD Enriched Brain Co-Expression Networks Classified by *APOE* Genotype and AD Status

Weighted gene co-expression network analysis (WGCNA) was conducted separately in AD cases and controls within each *APOE* genotype groups using data from the largest brain sample (ROSMAP) (**Supplementary Table 1**). Twenty-three gene co-expression networks, each enriched with differentially expressed genes (**Supplementary Table 2**) and AD risk genes established by large genome-wide association studies (GWAS) [1], were preserved in brain samples from the MAYO or FHS/BUADC datasets (**Supplementary Table 8**). We evaluated brain cell-type enrichment derived from analysis of single-nuclei RNA sequencing data for these 23 networks (**Supplementary Figures 6 and 7**). Eleven networks (notably not M01) were enriched in both inhibitory and excitatory neurons, and network M18 was exclusively enriched in these two cell-types. Four networks (M3, M6, M12, and M19) representing both AD cases and controls, as well as the *APOE* ε2/ε3 and ε3/ε4 genotype groups, were enriched in astrocytes, microglia, oligodendrocytes and oligodendrocyte progenitor cells (OPCs). Networks M11 and M15 were exclusively enriched in OPCs (**Fig. 2A and Supplementary Table 9**).

**Fig. 2.**
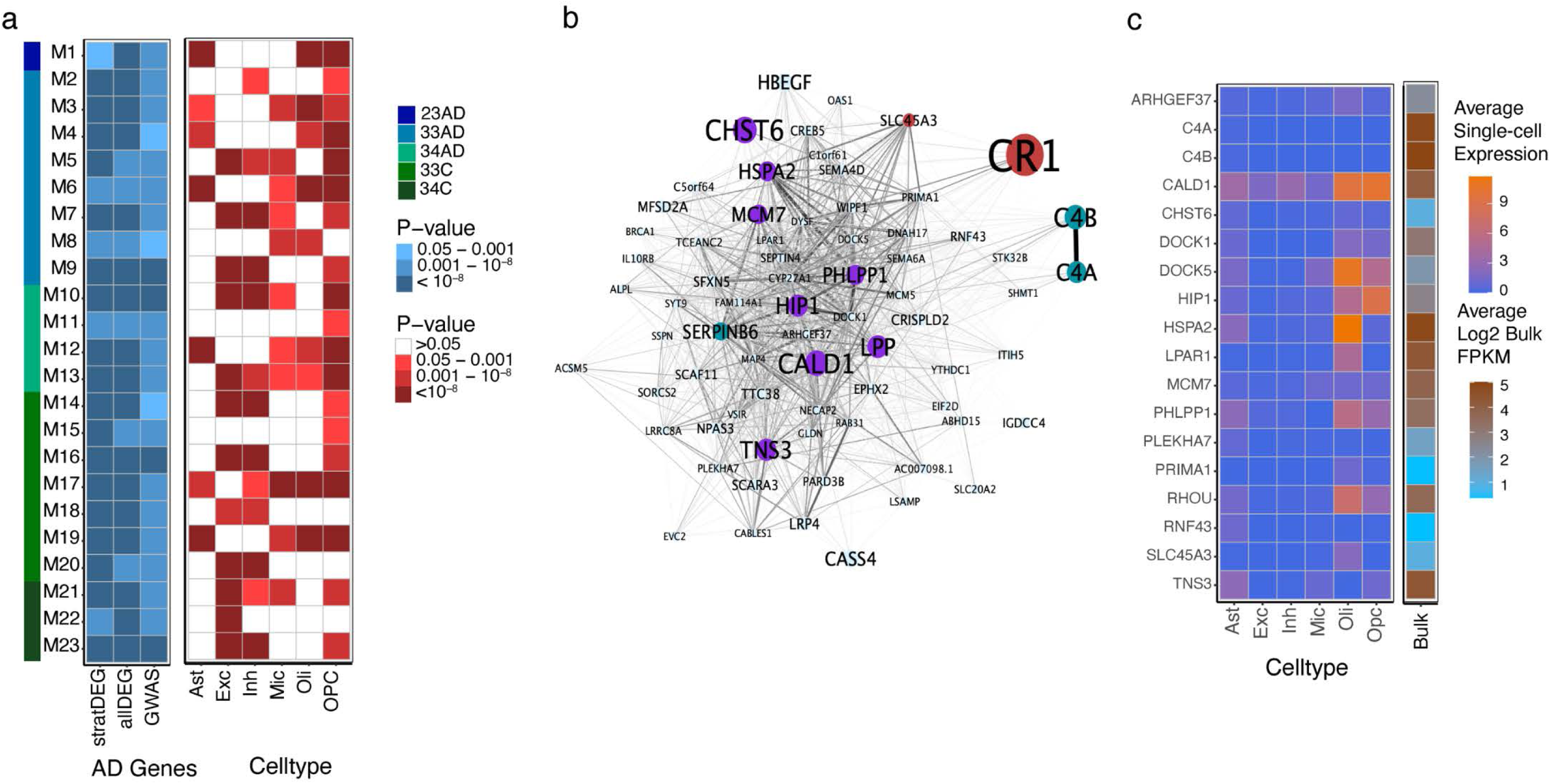
AD-related Gene Co-Expression Networks in Brain. **(a)** Heatmaps depicting association of co-expressed gene networks (modules M1-M23) derived from analysis within a subgroup defined by *APOE* genotype and AD status according to the color scheme shown on the right. The far-left vertical bar, the left blue-shaded panel, and the right red-shaded panel represents networks enriched in different *APOE* genotype subgroups, in differentially expressed genes between AD and control subjects, and in particular cell types, respectively. The AD gene heatmap showed enrichment of each gene co-expression network with genes that are differentially expressed in (1) the corresponding *APOE* genotype subgroup (stratDEG) or (2) the entire sample (allDEG), or with AD risk genes established by GWAS. The cell-type heatmap shows enrichment of each gene co-expression network for astrocytes (Ast), excitatory neurons (Exc), inhibitory neurons (Inh), microglia (Mic), oligodendrocytes (Oli), and oligodendrocyte progenitor cells (OPC). All enrichment p-values are Bonferroni corrected. **(b)** Gene co-expression network (M01) that was derived using WGCNA from analysis of *APOE* ε2/ε3 subjects with AD. Differentially expressed genes in the *APOE* ε2/ε3 genotype group (p<0.01) and in the total sample (p<10^−6^) are highlighted in turquoise. Genes associated with AD risk in GWAS (p<10^−3^) and differentially expressed in the total sample (p<10^−6^) are highlighted in purple. Genes associated with AD risk at the genome-wide significance level and differentially expressed in the total sample (p<10^−6^) as well as in the *APOE* ε2/ε3 genotype group (p<0.01) are highlighted in red. The size of each node inversely corresponds with the p-value supporting the association of the gene with AD. **(c)** Heatmaps showing the average expression of genes in ROSMAP subjects across cell-types calculated from analysis of single-nuclei RNA-seq data and in the bulk RNA-seq data from subjects overlapping the single-cell RNA-seq dataset. Genes are members of the M01 co-expression network whose expression was nominally associated (p<0.05) with plaque score and Braak stage. *C5orf64* was excluded as it did not occur in the single-cell expression dataset.

The M01 network specific to ε2/ε3 AD cases contained several complement pathway genes including *C4A, C4B*, and C3b/C4b Receptor 1 (*CR1*) (**Fig. 2B)**. No other networks contained all three of these genes suggesting that this network may be specific to AD subjects with the ε2/ε3 genotype. This ε2/ε3-AD network contained 674 genes of which 96 (10.2%) were differentially expressed in the total sample (Bonferroni corrected enrichment p-value [ENR-p]=4.3×10^−32^), eight (1.2%) were differentially expressed among ε2/ε3 subjects (ENR-p=0.05), and 66 (9.8%) were significantly associated with AD risk (ENR-p=1.2×10^−5^) (**Fig. 2A and Supplementary Table 8**). The ε2/ε3-AD network was also significantly enriched in different cell types including astrocytes (ENR-p=5.2×10^−59^), oligodendrocytes (ENR-p=9.1×10^−57^), and OPCs (ENR-p=6.8×10^−44^) (**Fig. 2A** and **Supplemental Table 9**). Of the 154 genes enriched uniquely in the ε2/ε3-AD network, expression of 19 was nominally associated (p<0.05) with Braak Stage and plaque density including *C4A* and *C4B* (**Supplementary Table 10**). Ten of these 19 genes were expressed in oligodendrocytes, astrocytes, and/or OPCs and four of these genes including *HSPA2, LPAR1, DOCK1*, and *PHLPP1* were significantly (FDR adjusted p<0.05) differentially expressed between AD and control oligodendrocytes (**Fig. 2C, Supplemental Figure 8 and Supplemental Table 11**). *HPSA2* and *DOCK1* were transcriptome-wide significant DEGs in the total sample, and differential expression of *HSPA2* was nominally significant in the ε2/ε3 group (p=0.05) (**Supplementary Table 2**). None of these 10 genes were differentially expressed between AD and control astrocytes and OPCs (**Supplemental Table 11**).

### Association of C4A/B and HSPA2 Expression with AD-Related Protein Levels

We tested association of expression of eight genes from the four top-ranked DEGs among *APOE* ε2/ε3 subjects (**Table 1**) and the top-ranked genes in the ε2/ε3-AD network (**Supplemental Figure 8**) with immunoassay measures of pTau181/tTau ratio, pTau231/tTau ratio, Aβ42, and postsynaptic density protein 95 (PSD95) as well as with levels of C4A and C4B proteins in FHS/BUADC brain tissue (**Supplementary Table 12)**. Expression of *C4A, C4B, GFAP, NPNT, HSPA2*, and *PHLPP*1 was significantly associated with pTau231/tTau at the multiple test correction threshold of p<6.3×10^−3^ (**Fig. 3A and Supplementary Table 13**), and results for *C4A, C4B*, and *HSPA2* surpassed an even more stringent threshold of P<10^−4^ (**Fig. 3B and Table 2**). *HSPA2* expression was significantly associated with C4B protein level after multiple testing correction (P=6.1×10^−3^) (**Fig. 4**). Expression of all eight genes in this group was not associated with Aβ_42,_ PSD95 or pTau181/tTau ratio after multiple testing correction.

**Table 2.** **Association of expression of co-expressed genes in *APOE ε2/ε3* AD cases with levels of AD-related traits and C4 subunit proteins**

| | Total Sample | | | *APOE* $\epsilon 2/\epsilon 3$ | | | *APOE* $\epsilon 3/\epsilon 3$ | | | *APOE* $\epsilon 3/\epsilon 4$ | | |
| --- | --- | --- | --- | --- | --- | --- | --- | --- | --- | --- | --- | --- |
| | $\beta$ | SE | P-value | $\beta$ | SE | P-value | $\beta$ | SE | P-value | $\beta$ | SE | P-value |
| **pTau231/tTau:** |  |  |  |  |  |  |  |  |  |  |  |  |
| *C4A* | 0.22 | 0.05 | 5.1x10<sup>-5</sup> | 0.21 | 0.10 | 0.04 | 0.18 | 0.08 | 0.02 | 0.34 | 0.12 | 6.4x10<sup>-3</sup> |
| *C4B* | 0.21 | 0.05 | 7.0x10<sup>-5</sup> | 0.19 | 0.10 | 0.06 | 0.18 | 0.08 | 0.02 | 0.34 | 0.11 | 5.4x10<sup>-3</sup> |
| *HSPA2* | 0.30 | 0.06 | 4.3x10<sup>-6</sup> | 0.31 | 0.13 | 0.02 | 0.20 | 0.10 | 0.04 | 0.41 | 0.14 | 6.1x10<sup>-3</sup> |
| **PSD95:** |  |  |  |  |  |  |  |  |  |  |  |  |
| *C4A* | -0.10 | 0.06 | 0.08 | -0.17 | 0.12 | 0.16 | -0.06 | 0.09 | 0.51 | -0.02 | 0.13 | 0.86 |
| *C4B* | -0.10 | 0.06 | 0.08 | -0.15 | 0.11 | 0.20 | -0.05 | 0.08 | 0.55 | -0.06 | 0.13 | 0.68 |
| *HSPA2* | -0.10 | 0.07 | 0.17 | -0.21 | 0.15 | 0.17 | -0.05 | 0.10 | 0.63 | -0.02 | 0.16 | 0.90 |
| **C4B:** |  |  |  |  |  |  |  |  |  |  |  |  |
| *C4A* | 0.06 | 0.05 | 0.33 | 0.14 | 0.13 | 0.29 | 0.01 | 0.09 | 0.90 | 0.19 | 0.14 | 0.19 |
| *C4B* | 0.05 | 0.05 | 0.43 | 0.14 | 0.13 | 0.26 | -0.01 | 0.09 | 0.88 | 0.19 | 0.14 | 0.18 |
| *HSPA2* | 0.20 | 0.07 | 6.1x10<sup>-3</sup> | 0.30 | 0.17 | 0.09 | 0.21 | 0.11 | 0.06 | 0.32 | 0.16 | 0.06 |

$\beta$ = effect size; SE = standard error

**Fig. 3.**
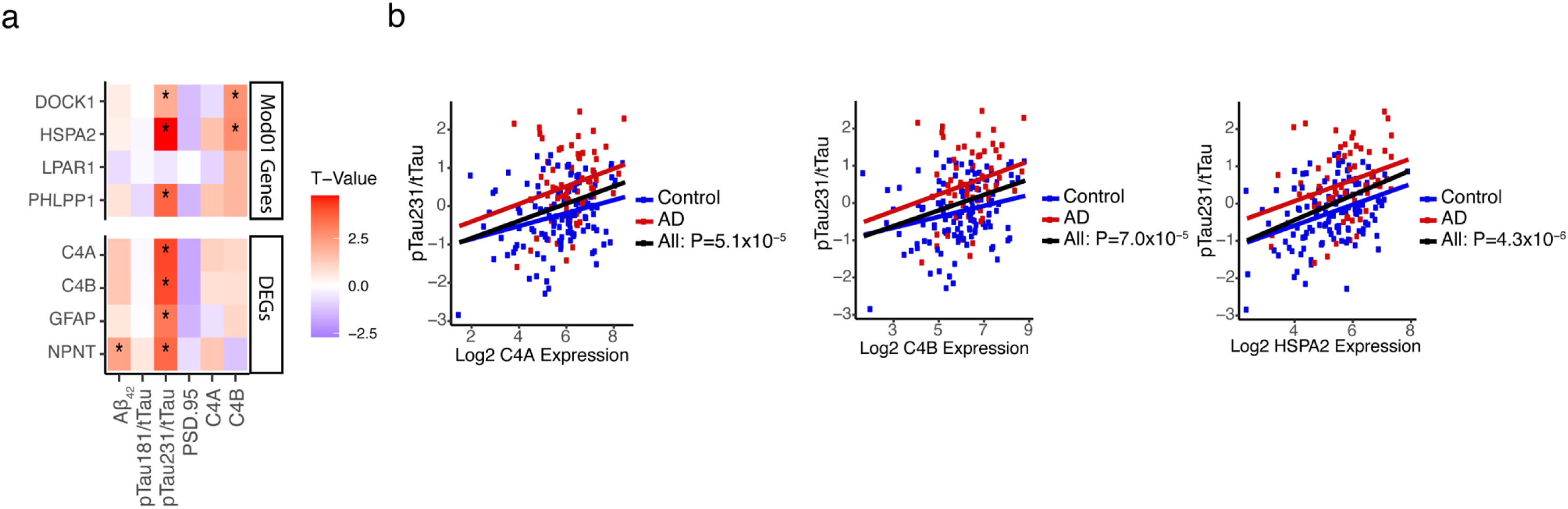
Association of differentially expressed genes among *APOE* ε2 carriers with AD-related proteins. **(a)** Heatmap showing the association of gene expression with and AD-related proteins. Genes shown are differentially expressed in the total sample (P<10^−6^) and among *APOE* ε2/ε3 subjects (P<10^−3^), or were selected from the M01 network and are significantly differentially expressed between AD and control oligodendrocytes. Nominally significant (P<0.05) associations are marked by an asterisk. **(b)** Scatterplots showing association of expression of genes in *APOE* ε2/ε3 co-expression network (*C4A, C4B*, and *HSPA2*) with rank-transformed pTau231/tTau ratio. Coordinates for each subject are shown as color-coded dots (red=AD, blue=controls) and their correlation across subjects (within AD cases and control groups, and for the total group) is represented by fitted solid lines (black=combined sample).

**Fig 4.**
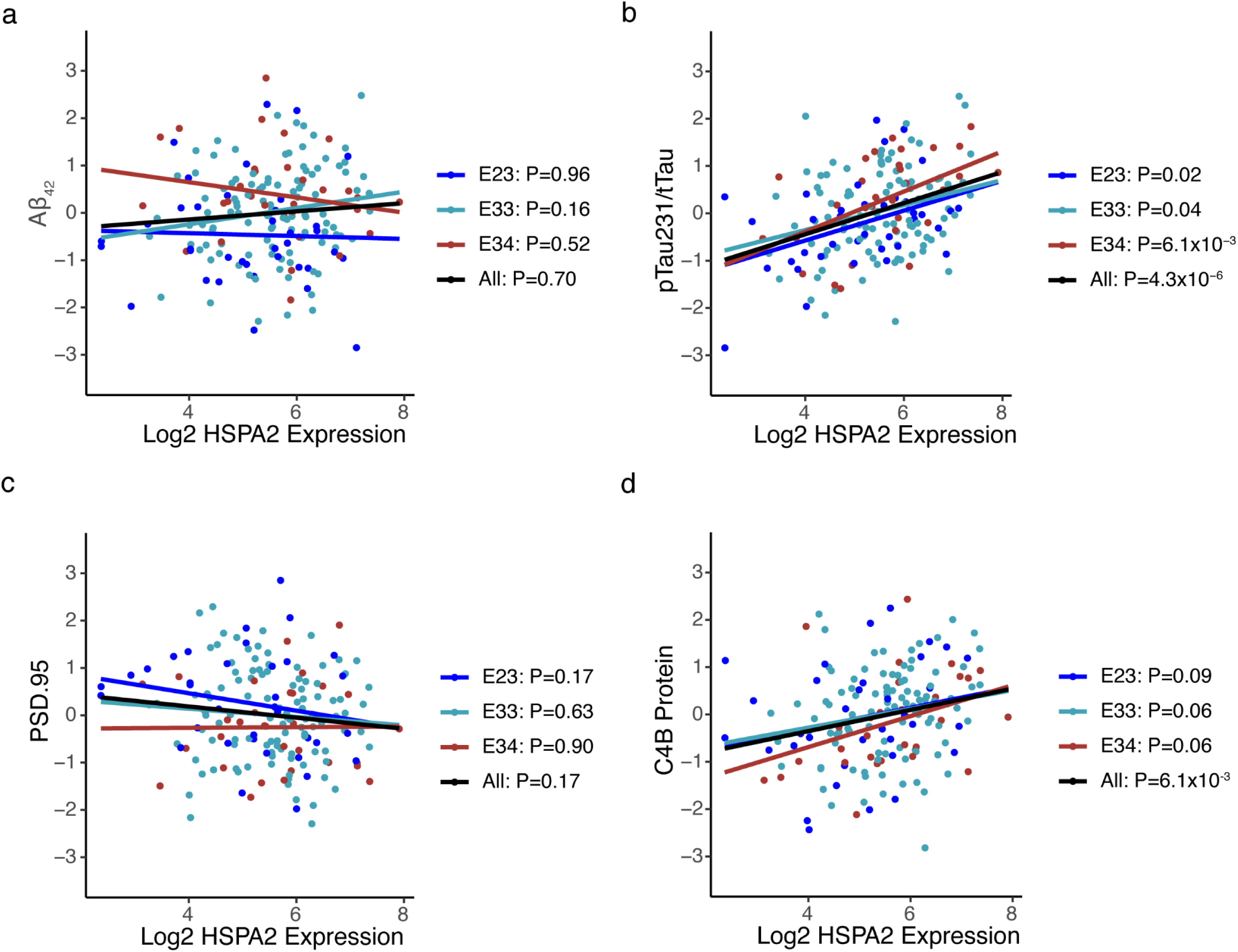
Association of *HSPA2* gene expression with AD-related proteins. Scatterplots showing association of *HSPA2* expression with rank-transformed **(a)** Aβ_42_ level, **(b)** pTau231/tTau ratio, **(c)** PSD95, and **(d)** C4B protein level. Coordinates are shown as color-coded dots by *APOE* genotype (ε2/ε3=dark blue, ε3/ε3=light blue, ε3/ε4=red) and their correlation across subjects (within *APOE* subgroups and for the total group) is represented by fitted solid lines (black=combined sample).

## Discussion

The primary purpose of this study was to discern genes and biological pathways that may have a role in the mechanism underlying the protective effect of *APOE* ε2 against AD. We identified 11 genes that were differentially expressed between AD cases and controls within *APOE* ε2/ε3, ε3/ε3, and ε3/ε4 genotype groups, suggesting that their influence on AD risk is likely not specific to any *APOE* genotype. We identified transcriptome-wide significant differential expression of three genes, *C4A, C4B*, and *GFAP*, for which the evidence was strongest among *APOE* ε2/ε3 individuals. We also identified 23 brain cell-type specific networks that are enriched for significant AD-associated genes and DEGs and characterized by unique biological pathways. One of these networks, M01, was specific to AD cases with the *APOE* ε2/ε3 genotype and contained multiple genes in classical complement pathway including *C4A, C4B*, and *CR1*. Expression of *C4A, C4B*, and *HSPA2* was significantly associated with amyloid plaque and neurofibrillary tangle density, as well as with the ratio of phosphorylated tau at protein position 231 to total Tau (pTau231/tTau). Expression of *HSPA2*, a significant DEG between AD and control oligodendrocytes, was significantly associated with the level of C4B protein. These findings suggest that the co-regulated top-ranked genes in the *APOE* ε2/ε3-AD network are likely involved in regulation of classical complement activation and tau phosphorylation.

The role of neuroinflammation in AD has become increasingly important, especially after repeated disappointing results of drugs targeting the processing and toxic isoforms of Aβ to ameliorate symptoms or halt progression of AD [44]. The classical complement pathway has been consistently linked with neuroinflammation and neuroimmune response [45], but has garnered much less attention regarding its role in AD. We found that expression of two genes in this pathway, *C4A* and *C4B*, were significantly up-regulated in AD cases compared to controls regardless of *APOE* genotype, but the difference was larger and more significant among ε2/ε3 individuals despite the smaller sample size compared to other *APOE* genotypes. The astrocyte specific marker gene, *GFAP*, was also up-regulated most significantly in ε2/ε3 individuals. *GFAP* has previously been associated with the increase of reactive astrocytes in AD [46]. Reactive astrocytes have been shown to up-regulate the complement cascade as part of their neuroimmune response [47].

The complement pathway has recently been linked to *APOE* through binding with activated C1q creating a *APOE*-C1q complex [48]. *CR1*, one of the most well-established AD genes [49], was a member of the unique *APOE* ε2/ε3 AD co-expression network in our study which also contained *C4A* and *C4B*. CR1 is a known receptor for C1q as well as C4A and C4B, and can bind to C1q and C4B simultaneously [50]. ApoE can activate the complement pathway *in vitro* and cause deposition of C4B, but only in the presence of C1q [51], suggesting that C1q could possibly facilitate an ApoE/C4 interaction through CR1. Future studies are needed to confirm the binding interactions among members of this network, and determine whether this network is disrupted in the *APOE* genotype dependent manner and, if so, elucidate the nature of the interaction between the complement pathway and ε2.

The only significant co-expression network specific to *APOE* ε2/ε3 AD brains was enriched for astrocytes, oligodendrocytes, and OPCs. Astrocytes have been implicated with the classical complement pathway and general neuroimmune response, and can increase expression of complement components [47]. Oligodendrocytes can express complement component mRNAs, however, their role in the classical complement pathway is not well understood [52]. One study found complement component 4 exclusively co-localized with oligodendrocytes in APP transgenic mouse models for AD [53]. Among the significant DEGs genes in the *APOE* ε2/ε3-AD network, *HSPA2* was expressed predominantly in oligodendrocytes and is connected to the classical complement pathway via its association with C4B protein level in the brain. *HSPA2* was previously associated with AD risk using systems biology approaches for analysis of multi-omics data including network diffusion which integrates information from protein network analysis and GWAS [54], analysis of brain region co-expression networks [55], and Bayesian network analysis [56]. In addition, heat shock-related 70-kDa protein 2 (encoded by *HSPA2*) has been linked to myelination function and fast cognitive decline [57]. While AD is generally considered a gray matter disease [58], white matter abnormalities have been observed in AD including loss of myelination and inability of oligodendrocytes to repair myelin [59]. Further studies are required to understand the connection between oligodendrocyte-specific *HSPA2* expression and complement component 4 in AD.

Two other *APOE* ε2/ε3-AD network genes have also been linked to AD-related processes. *PHLPP1*, whose expression was specific to oligodendrocytes, OPCs, and astrocytes, encodes PH domain and leucine rich repeat protein phosphatase 1 which helps regulate protein kinase B (AKT) [60], and the dysregulation of AKT can cause reduced tau phosphorylation [61]. *DOCK1*, expressed only in oligodendrocytes, encodes Dock180 which has a role in dendritic spine morphogenesis and axon pathfinding [62]. *C4A* and *C4B* were not expressed in any cell-types in our single-nuclei RNA sequencing data, perhaps because *C4A* and *C4B* mRNAs localized outside the nucleus and thus their expression was not detected in our analysis of snRNA-seq data.

Six of the eight top-ranked genes in the *APOE* ε2/ε3-AD network were significantly associated with the pTau231/tTau ratio, but not with the pTtau181/tTau ratio. This observation is consistent with a previous study showing that pTau231/tTau may be a better indicator of AD-related tau mechanisms [63]. None of the top-ranked genes in this network were associated with Aβ_42_ level, suggesting that this network is more involved in processes related to tau but not Aβ. The connection of ε2 to tau is supported by evidence that ε2 is associated with increased tau in mice expressing *APOE* ε2/ε2 and with increased tau pathology in the brains of human tauopathy cases [64]. In a separate study (see the companion paper by Jun et al), we linked *PPP2CB*, a hub of an *APOE* ε2-related gene network associated with AD, to C4B protein level in brain. In addition, we demonstrated that expression of *PPP2CB*, which encodes one of the catalytic subunits of protein phosphatase 2A (PP2A), was significantly correlated with pTau231/tTau in human brain and in isogenic *APOE* human induced pluripotent stem cell (iPSC)-derived neurons co-cultured with astrocytes (Jun et al.), and dysfunction of PP2A promotes tau hyperphosphorylation [65]. In the current study, *C4A* and *C4B* expression was not associated with C4A and C4B protein concentration levels, however, this may be due to altered protein degradation and/or a variety of gene expression regulation factors [66].

Although our findings demonstrate that genes in the classical complement pathway have a role in AD likely through interaction with *APOE* ε2, mechanisms underlying the connection between the complement pathway and the protective effect of *APOE* ε2 against AD are still unclear. It is possible that reduced expression of complement components, such as *C4A* and *C4B*, in the presence of ε2 is neuroprotective, an idea consistent with separate evidence of neuroprotective effects of ε2 with respect to AD [67,68]. Complement pathway mRNAs are generally up-regulated in brain regions affected by AD [69]. Inhibition of several complement components, including C1q and C3aR, in tau mutation mouse models can cause attenuation of neurodegeneration especially synaptic loss [70,71]. Further studies are necessary to examine the potential neuroprotective effect of reduced mRNA levels of complement components in AD, especially in relationship to the *APOE* ε2 allele.

Our study has several limitations. First, the number of individuals with the *APOE* ε2 allele, was small and, hence, power for analyses specific to ε2 subjects was lower than for those with other *APOE* genotypes. There was also an unbalanced distribution of *APOE* ε3/ε4 AD and control individuals especially in the ROSMAP dataset. We were able to mitigate some of these issues through meta-analysis of results across datasets. However, there was insufficient power for interaction tests due to the relatively small sizes of some *APOE* genotype groups. Second, pooling results across all datasets may have led to inconsistent findings because transcriptomic data were derived from temporal cortex in the MAYO sample and prefrontal cortex in the ROSMAP and BUADC/FHS samples. However, the concern about brain region variability is low because both temporal and prefrontal cortex are both severely affected by AD [72,73]. Third, measures of plaque and tangle density were unavailable in the MAYO dataset and, thus, results involving these variables are based on a smaller sample. As a result, we were unable to conduct meta-analysis of all three datasets. Fourth, gene expression patterns and levels of AD-related proteins in brains from AD cases may be indicative of post-mortem changes unrelated to AD pathogenesis, noting also that RNA degradation occurs faster than protein degradation [74]. Fifth, although WGCNA is a powerful computational tool for identifying gene co-expression patterns, this method does not consider any biological implications when creating networks. Finally, single-nuclei RNA sequence analysis limits interpretation of expression findings to the nucleus instead of the whole cell. Taken together, these caveats emphasize the need for replication in independent samples and validation using other approaches.

In summary, our findings provide further evidence that the complement cascade, and the classical complement pathway in particular, has an important role in AD. Complement proteins including C4A and C4B, as well as HSPA2 protein that we linked to the complement pathway, may confer a neuroprotective effect against AD through interaction with *APOE* ε2.

## Data Availability

Publicly available RNA sequencing and neuropathological data were obtained from the CommonMind Consortium portal

http://www.synapse.org

## Acknowledgements

This study was supported by the National Institute on Aging (NIA) grants RF1-AG057519, R01-AG048927, P30-AG013846, U01-AG032984, U19-AG068753, U01-AG062602, U01-AG058654, RF1-AG054156, RF1-AG08122, RF1-AG054076, RF1-AG054199, R01-AG066429, and R01-AG054672, and by Framingham Heart Study contracts 75N92019D00031 and HHSN2682015000011. Collection of study data provided by the Rush Alzheimer’s Disease Center, Rush University Medical Center, Chicago was supported through funding by NIA grants P30-AG10161, R01-AG15819, R01-AG17917, R01-AG30146, R01-AG36836, U01-AG32984, U01-AG46152, and U01-AG61358, and funding from the Illinois Department of Public Health and the Translational Genomics Research Institute. Study data were also provided by Dr. Nilüfer Ertekin-Taner and Dr. Steven G. Younkin, Mayo Clinic, Jacksonville, FL using samples from the Mayo Clinic Study of Aging, the Mayo Clinic Alzheimer’s Disease Research Center, and the Mayo Clinic Brain Bank. Collection of these data was supported through funding by NIH grants P50-AG016574, R01-AG032990, U01-AG046139, R01-AG018023, U01-AG006576, U01-AG006786, R01-AG025711, R01-AG017216, R01-AG003949, and R01-NS080820, and by funding from the CurePSP Foundation and the Mayo Foundation.

## Author Contributions

R.P., L.A.F., and G.R.J. conceived study design. R.P. and G.R.J. conceived statistical analysis. R.P., J.H., J.C., and C.Z. performed data analyses. R.P., T.D.S. L.A.F., and G.R.J. wrote the manuscript. D.A.B. and R.A. provided data critical to the study. K.L.L., T.I., D.A.B., T.D.S., L.A.F., and G.R.J. provided expertise and edited the manuscript. G.M., W.X., and T.D.S. conceived and performed immunoassay experiments. L.A.F. and G.R.J. obtained funding for this project.

