## Supplemental Materials for "Integrative Brain Transcriptome Analysis Links Complement Component 4 and *HSPA2* to the *APOE* ε2 Protective Effect in Alzheimer Disease"

### Supplementary Table 1. Sample characteristics by source and *APOE* genotype

| Dataset | APOE Genotype | AD Cases | | | | | Controls | | | | |
|---|---|---|---|---|---|---|---|---|---|---|---|
| | | N | % Female | AAD | PMI | RIN | N | % Female | AAD | PMI | RIN |
| ROSMAP | ε2/ε2 | 0 | NA | NA | NA | NA | 5 | 60.0 | 92.8 ± 4.5 | 6.4 ± 2.4 | 6.2 ± 0.9 |
| | ε2/ε3 | 38 | 81.6 | 92.7 ± 6.7 | 7.8 ± 7.0 | 7.3 ± 1.0 | 44 | 65.9 | 88.4 ± 8.5 | 6.2 ± 3.8 | 7.2 ± 1.2 |
| | ε3/ε3 | 212 | 65.6 | 90.2 ± 5.9 | 7.8 ± 5.3 | 7.0 ± 1.0 | 168 | 58.9 | 86.5 ± 6.9 | 7.2 ± 4.3 | 7.3 ± 1.0 |
| | ε2/ε4 | 12 | 58.3 | 89.9 ± 3.4 | 6.8 ± 2.5 | 7.1 ± 1.0 | 4 | 50.0 | 89.0 ± 5.8 | 6.5 ± 5.5 | 6.9 ± 1.3 |
| | ε3/ε4 | 114 | 65.8 | 88.7 ± 5.6 | 7.1 ± 4.4 | 7.0 ± 0.9 | 25 | 56.0 | 85.7 ± 7.8 | 7.3 ± 5.0 | 7.2 ± 1.1 |
| | ε4/ε4 | 4 | 50.0 | 89.9 ± 1.3 | 13.0 ± 7.9 | 6.7 ± 0.5 | 1 | 100 | 85.4 | 5.8 | 5.9 |
| | TOTAL | 380 | 66.8 | 90.0 ± 5.9 | 7.6 ± 5.2 | 7.0 ± 0.9 | 247 | 59.9 | 86.9 ± 7.3 | 7.0 ± 4.2 | 7.2 ± 1.0 |
| MAYO | ε2/2 | 0 | NA | NA | NA | NA | 0 | NA | NA | NA | NA |
| | ε2/3 | 4 | 50.0 | 85.8 ± 6.6 | NA | 8.9 ± 0.2 | 12 | 41.7 | 81.4 ± 10.0 | NA | 7.7 ± 1.0 |
| | ε3/3 | 35 | 54.3 | 83.7 ± 8.0 | NA | 8.6 ± 0.5 | 58 | 48.3 | 82.4 ± 9.1 | NA | 7.6 ± 1.1 |
| | ε2/4 | 0 | NA | NA | NA | NA | 1 | 100 | 90 | NA | 6.7 |
| | ε3/4 | 36 | 63.9 | 82.9 ± 6.8 | NA | 8.5 ± 0.6 | 8 | 50.0 | 84.1 ± 5.4 | NA | 7.6 ± 1.0 |
| | ε4/4 | 7 | 71.4 | 74.2 ± 6.0 | NA | 8.5 ± 0.5 | 1 | 100 | 86.0 | NA | 5.9 |
| | TOTAL | 82 | 59.8 | 82.7 ± 7.6 | NA | 8.6 ± 0.5 | 80 | 48.8 | 82.6 ± 8.8 | NA | 7.6 ± 1.0 |
| FHS/ BUADC | ε2/2 | 1 | 100 | 68.0 | NA | 4.5 | 2 | 50.0 | 77.5 ± 20.5 | NA | 7.3 ± 1.1 |
| | ε2/3 | 6 | 66.7 | 84.8 ± 18.5 | NA | 5.8 ± 1.4 | 31 | 61.3 | 82.0 ± 11.2 | NA | 6.1 ± 1.4 |
| | ε3/3 | 33 | 33.3 | 90.5 ± 8.0 | NA | 6.9 ± 1.3 | 76 | 43.4 | 87.2 ± 8.6 | NA | 6.8 ± 1.3 |
| | ε2/4 | 3 | 66.7 | 80.7 ± 4.2 | NA | 5.5 ± 1.3 | 8 | 62.5 | 76.0 ± 11.0 | NA | 6.2 ± 1.1 |
| | ε3/4 | 19 | 42.1 | 89.8 ± 6.4 | NA | 6.6 ± 1.2 | 12 | 58.3 | 82.8 ± 10.4 | NA | 7.0 ± 1.2 |
| | ε4/4 | 2 | 50.0 | 79.0 ± 18.3 | NA | 6.7 ± 0.6 | 0 | NA | NA | NA | NA |
| | TOTAL | 64 | 42.2 | 88.6 ± 9.7 | NA | 6.6 ± 1.3 | 129 | 50.4 | 84.7 ± 10.2 | NA | 6.6 ± 1.3 |

N: sample size; AAD: Age at death; PMI: Post-mortem interval; RIN: RNA integrity number. NA: not available. Values for AAD, PMI and RIN expressed as mean ± standard deviation. A diagnosis of definite AD was established according to the National Institute of Aging (NIA) Reagan criteria.

**Supplementary Table 3.** Top-ranked differentially expressed genes in the *APOE* ε2/ε3 group

| Gene | ROSMAP | | MAYO | | FHS/BUADC | | Total | |
|---|---|---|---|---|---|---|---|---|
| | **LogFC** | **P-value** | **LogFC** | **P-value** | **LogFC** | **P-value** | **Z-score** | **P-value** |
| *C4A* | 1.05 | $6.0 \times 10^{-5}$ | 1.42 | 0.05 | 0.30 | 0.66 | 4.28 | $1.9 \times 10^{-5}$ |
| *C4B* | 1.00 | $7.3 \times 10^{-5}$ | 0.98 | 0.12 | 0.30 | 0.67 | 4.10 | $4.2 \times 10^{-5}$ |
| *GFAP* | 0.89 | $1.6 \times 10^{-5}$ | 0.86 | 0.15 | -0.12 | 0.81 | 4.12 | $3.8 \times 10^{-5}$ |
| *NPNT* | 0.40 | 0.02 | 1.03 | $2.1 \times 10^{-3}$ | 0.46 | 0.18 | 3.65 | $2.6 \times 10^{-4}$ |

Log2 fold change of expression levels (LogFC) between AD cases and controls was calculated within each dataset and the results were combined by meta-analysis.

**Supplementary Table 4.** Top-ranked differentially expressed genes ($p<0.01$) in each *APOE* genotype group in the combined ROSMAP, MAYO, and FHS/BUADC sample

| Gene | *APOE* ε2/ε3 | | *APOE* ε3/ε3 | | APOE ε3/ε4 | | Total | |
|---|---|---|---|---|---|---|---|---|
| | Z-score | P-value | Z-score | P-value | Z-score | P-value | Z-score | P-value |
| *C1QTNF5* | 3.18 | $1.5x10^{-3}$ | 6.02 | $1.8x10^{-9}$ | 4.08 | $4.6x10^{-5}$ | 8.45 | $2.8x10^{-17}$ |
| *SLC6A9* | 2.73 | $6.3x10^{-3}$ | 5.18 | $2.2x10^{-7}$ | 3.50 | $4.7x10^{-4}$ | 8.22 | $2.1x10^{-16}$ |
| *S100A4* | 2.65 | $8.0x10^{-3}$ | 4.42 | $9.9x10^{-6}$ | 3.54 | $4.1x10^{-4}$ | 7.59 | $3.3x10^{-14}$ |
| *ADAMTS2* | 2.65 | $8.1x10^{-3}$ | 4.10 | $4.1x10^{-5}$ | 4.20 | $1.7x10^{-4}$ | 7.22 | $5.1x10^{-13}$ |
| *MUC1* | 3.12 | $1.8x10^{-3}$ | 3.94 | $8.0x10^{-5}$ | 4.04 | $5.3x10^{-5}$ | 7.02 | $2.6x10^{-12}$ |
| *CHD4* | 2.89 | $3.9x10^{-3}$ | 4.21 | $2.6x10^{-5}$ | 4.69 | $2.8x10^{-6}$ | 6.95 | $3.7x10^{-12}$ |
| *SH3PXD2B* | 3.06 | $2.2x10^{-3}$ | 3.70 | $2.2x10^{-4}$ | 3.62 | $2.9x10^{-4}$ | 6.40 | $1.6x10^{-10}$ |
| *DUSP4* | -2.96 | $3.0x10^{-3}$ | -3.05 | $2.3x10^{-3}$ | -2.69 | $7.1x10^{-3}$ | -5.82 | $6.0x10^{-9}$ |
| *SLC25A26* | -2.67 | $7.5x10^{-3}$ | -3.32 | $9.0x10^{-4}$ | -3.02 | $2.6x10^{-3}$ | -5.77 | $8.1x10^{-9}$ |
| *RRNAD1* | 2.81 | $4.9x10^{-3}$ | 3.03 | $2.5x10^{-3}$ | 2.70 | $6.9x10^{-3}$ | 5.44 | $5.3x10^{-8}$ |
| *RIMS1* | -2.82 | $4.8x10^{-3}$ | -3.02 | $2.5x10^{-3}$ | -3.27 | $1.1x10^{-3}$ | -5.17 | $2.3x10^{-7}$ |

Results were generated from bulk RNA sequence data and combined across data sets by meta-analysis of differential expression between AD and control brains.

**Supplementary Table 5.** Distribution of neurofibrillary tangles (quantified by Braak Stage) and neuritic plaques (quantified by CERAD plaque score) by dataset and *APOE* genotype

| Dataset | Subgroup | Braak Stage | | Plaque Score | |
|---|---|---|---|---|---|
| | | N | Mean+SD | N | Mean+SD |
| ROSMAP | *APOE* ε2/ε3 | 71 | -0.43 ± 0.8 | 71 | 0.55 ± 0.9 |
| | *APOE* ε3/ε3 | 349 | -0.20 ± 1.0 | 349 | 0.18 ± 1.0 |
| | *APOE* ε3/ε4 | 125 | 0.36 ± 0.9 | 125 | -0.45 ± 0.9 |
| | Total | 568 | -0.10 ± 1.0 | 568 | 0.08 ± 1.0 |
| FHS/BUDADC | *APOE* ε2/ε3 | 15 | -0.55 ± 0.7 | 13 | 0.52 ± 0.8 |
| | *APOE* ε3/ε3 | 99 | -0.09 ± 1.0 | 77 | 0.13 ± 1.0 |
| | *APOE* ε3/ε4 | 30 | 0.47 ± 1.0 | 26 | -0.45 ± 0.9 |
| | Total | 151 | -0.03 ± 1.0 | 122 | 0.06 ± 1.0 |

N= sample size; SD= standard deviation

Plaque (CERAD) scores reflect the density of neuritic plaques: 1 (definite), 2 (probable), 3 (possible, and 4 (none); Braak stages represent the spatial distribution of neurofibrillary tangles: 0 (none), I-II (transentorhinal and entorhinal cortex), III-IV (hippocampal and neighboring limbic areas), and V-VI (neocortical areas). All traits were rank-transformed for age of death and sex.

**Supplementary Table 6.** Association of expression of top-ranked differentially expressed genes among *APOE* ε2/ε3 subjects with neurofibrillary tangles (Braak stage) and neuritic plaques (plaque score) in brains from AD cases and controls in the ROSMAP and FHS/BUADC datasets.

| **Gene** | **Braak Stage** | | | | | | **Plaque Score** | | | | | |
|---|---|---|---|---|---|---|---|---|---|---|---|---|
| | **ROSMAP** | | **FHS/BUADC** | | **Meta-Analysis** | | **ROSMAP** | | **FHS/BUADC** | | **Meta-Analysis** | |
| | **β** | **P-value** | **β** | **P-value** | **β** | **P-value** | **β** | **P-value** | **β** | **P-value** | **β** | **P-value** |
| *C4A* | 0.15 | $5.4x10^{-4}$ | 0.09 | 0.36 | 0.14 | $3.7x10^{-4}$ | -0.13 | $2.8x10^{-3}$ | -0.14 | 0.23 | -0.13 | $1.2x10^{-3}$ |
| *C4B* | 0.16 | $2.6x10^{-4}$ | 0.13 | 0.21 | 0.16 | $1.1x10^{-4}$ | -0.16 | $5.3x10^{-4}$ | -0.19 | 0.11 | -0.16 | $1.3x10^{-4}$ |
| *GFAP* | 0.21 | $5.4x10^{-6}$ | 0.08 | 0.30 | 0.18 | $9.1x10^{-6}$ | -0.18 | $1.8x10^{-4}$ | -0.13 | 0.13 | -0.17 | $5.5x10^{-5}$ |
| *NPNT* | 0.32 | $1.4x10^{-6}$ | 0.09 | 0.11 | 0.19 | $1.2x10^{-5}$ | -0.27 | $7.4x10^{-5}$ | -0.17 | $6.9x10^{-3}$ | -0.22 | $2.2x10^{-6}$ |

Expression differences (β estimate represent the magnitude of effect) for each trait were calculated using a linear regression model separately in each dataset and the results were combined by meta-analysis.

**Supplementary Table 7.** Association of expression of top-ranked differentially expressed genes among *APOE* ε2/ε3 subjects with neurofibrillary tangles (Braak stage) and neuritic plaques (plaque score) in the ROSMAP dataset

| Gene | *APOE* genotype | Braak Stage | | Plaque Score | |
|---|---|---|---|---|---|
| | | β | P-value | β | P-value |
| *C4A* | ε2/ε3 | 0.22 | $9.8x10^{-3}$ | -0.27 | $5.2x10^{-3}$ |
| | ε3/ε3 | 0.15 | $8.9x10^{-3}$ | -0.05 | 0.35 |
| | ε3/ε4 | 0.02 | 0.85 | -0.16 | 0.10 |
| | Total | 0.15 | $5.4x10^{-4}$ | -0.13 | $2.8x10^{-3}$ |
| *C4B* | ε2/ε3 | 0.20 | 0.03 | -0.31 | $2.2x10^{-3}$ |
| | ε3/ε3 | 0.17 | $3.3x10^{-3}$ | -0.08 | 0.13 |
| | ε3/ε4 | 0.05 | 0.63 | -0.18 | 0.08 |
| | Total | 0.16 | $2.6x10^{-4}$ | -0.16 | $5.3x10^{-4}$ |
| *GFAP* | ε2/ε3 | 0.26 | $7.1x10^{-3}$ | -0.33 | $2.5x10^{-3}$ |
| | ε3/ε3 | 0.18 | $3.4x10^{-3}$ | -0.09 | 0.15 |
| | ε3/ε4 | 0.12 | 0.22 | -0.09 | 0.34 |
| | Total | 0.21 | $5.4x10^{-6}$ | -0.18 | $1.8x10^{-4}$ |
| *NPNT* | ε2/ε3 | 0.16 | 0.29 | -0.25 | 0.14 |
| | ε3/ε3 | 0.38 | $5.9x10^{-6}$ | -0.28 | $5.7x10^{-4}$ |
| | ε3/ε4 | 0.12 | 0.43 | 0.04 | 0.79 |
| | Total | 0.32 | $1.4x10^{-6}$ | -0.27 | $7.4x10^{-5}$ |

Expression differences (β estimate represent the magnitude of effect) for each trait were calculated using a linear regression model separately in each dataset and the results were combined by meta-analysis.

**Supplementary Table 8**. Robust gene co-expression networks derived from the ROSMAP sample using various sources of AD-associated seed genes that are preserved in MAYO and/or FHS/BUADC datasets

| Mod | Total Genes | *APOE* and AD Status Subgroup | Preservation Score[1] | | Source of Genes Used to Seed Co-expression Networks | | | | | |
|---|---|---|---|---|---|---|---|---|---|---|
| | | | | | DEGs in Total Sample[2] | | DEGs in *APOE* Subgroup[3] | | AD GWAS[4] | |
| | | | MAYO | FHS/BUADC | # Genes | ENR-P | # Genes | ENR-P | # Genes | ENR-P |
| M01 | 674 | 23ad | 28.2 | -0.8 | 96 | $4.3x10^{-32}$ | 8 | 0.05 | 66 | $1.2x10^{-5}$ |
| M02 | 1501 | 33ad | 28.7 | 25.1 | 108 | $1.2x10^{-16}$ | 430 | $7.1x10^{-86}$ | 111 | $8.4x10^{-6}$ |
| M03 | 112 | 33ad | 30.7 | 5.7 | 23 | $7.0x10^{-12}$ | 56 | $1.1x10^{-23}$ | 18 | $1.6x10^{-4}$ |
| M04 | 203 | 33ad | 35.8 | 7.0 | 71 | $3.5x10^{-55}$ | 121 | $3.3x10^{-63}$ | 21 | 0.02 |
| M05 | 449 | 33ad | 58.9 | 31.7 | 39 | $7.3x10^{-8}$ | 222 | $1.8x10^{-94}$ | 45 | $2.8x10^{-5}$ |
| M06 | 495 | 33ad | 61.2 | 23.4 | 35 | $7.6x10^{-5}$ | 90 | $1.6x10^{-5}$ | 47 | $7.2x10^{-5}$ |
| M07 | 774 | 33ad | 120.6 | 37.5 | 126 | $7.6x10^{-56}$ | 472 | $8.1x10^{-260}$ | 73 | $1.1x10^{-7}$ |
| M08 | 247 | 33ad | 16.9 | 6.4 | 59 | $7.0x10^{-4}$ | 59 | $1.1x10^{-7}$ | 24 | 0.02 |
| M09 | 546 | 33ad | 205.9 | 39.0 | 84 | $8.2x10^{-35}$ | 379 | $3.0x10^{-238}$ | 78 | $6.7x10^{-18}$ |
| M10 | 2105 | 34ad | 91.2 | 46.3 | 247 | $5.9x10^{-77}$ | 350 | $3.6x10^{-108}$ | 210 | $6.8x10^{-24}$ |
| M11 | 1242 | 34ad | 14.3 | 23.7 | 69 | $1.3x10^{-4}$ | 102 | $1.4x10^{-7}$ | 99 | $1.8x10^{-5}$ |
| M12 | 407 | 34ad | 42.6 | 17.6 | 152 | $1.5x10^{-121}$ | 131 | $6.9x10^{-74}$ | 45 | $1.0x10^{-5}$ |
| M13 | 491 | 34ad | 86.0 | 39.0 | 76 | $9.4x10^{-30}$ | 191 | $1.1x10^{-126}$ | 51 | $1.1x10^{-5}$ |
| M14 | 314 | 33c | 94.6 | 31.5 | 71 | $1.0x10^{-39}$ | 236 | $9.5x10^{-157}$ | 32 | $1.7x10^{-3}$ |
| M15 | 707 | 33c | 16.4 | 28.1 | 44 | $2.7x10^{-4}$ | 161 | $6.2x10^{-18}$ | 59 | $5.7x10^{-4}$ |
| M16 | 779 | 33c | 166.7 | 51.4 | 128 | $1.4x10^{-55}$ | 521 | $1.3x10^{-310}$ | 108 | $6.2x10^{-23}$ |
| M17 | 198 | 33c | 23.3 | 7.6 | 32 | $4.1x10^{-13}$ | 84 | $4.4x10^{-28}$ | 27 | $3.1x10^{-5}$ |
| M18 | 910 | 33c | 30.6 | 39.5 | 72 | $4.0x10^{-12}$ | 335 | $2.0x10^{-94}$ | 80 | $1.7x10^{-6}$ |
| M19 | 599 | 33c | 41.4 | 24.8 | 70 | $9,6x10^{-21}$ | 148 | $6.1x10^{-20}$ | 59 | $2.5x10^{-6}$ |
| M20 | 2234 | 33c | 43.9 | 45.0 | 103 | $4.0x10^{-4}$ | 512 | $3.7x10^{-61}$ | 158 | $2.5x10^{-6}$ |
| M21 | 320 | 34c | 51.4 | 19.0 | 50 | $1.1x10^{-17}$ | 136 | $1.9x10^{-89}$ | 38 | $9.1x10^{-5}$ |
| M22 | 254 | 34c | 34.3 | 19.4 | 37 | $5.2x10^{-12}$ | 35 | $2.4x10^{-06}$ | 38 | $2.0x10^{-7}$ |
| M23 | 782 | 34c | 90.1 | 36.0 | 144 | $3.3x10^{-62}$ | 162 | $1.1x10^{-54}$ | 91 | $2.0x10^{-11}$ |

ENR-P: Bonferroni corrected enrichment P-value; DEGs: differentially expressed genes.

[1] Preservation score: Z-summary statistic representing a network identified in the ROSMAP dataset across the MAYO and FHS/BUADC datasets. Interpretation of Z-summary statistic: < 2 = no preservation, 2-10 = moderate preservation, > 10 = high preservation.

[2] Includes DEGs in the total sample with $p<10^{-6}$

[3] Includes moderately significant DEGs in the corresponding *APOE* subgroup (p<0.01) that were also moderately significant in the total sample (p<0.01)

[4] Includes genes with AD association p-value $<10^{-3}$ large GWAS (Kunkle et al. Nature Genetics 2019; 51:414-430)

**Supplementary Table 9. Enrichment of highly preserved co-expression networks for brain cell type genes in the ROSMAP sample**

| Module | Total Genes | *APOE* and AD Status Subgroup | Enrichment P-value | | | | | |
|---|---|---|---|---|---|---|---|---|
| | | | Ast | Mic | Oli | OPC | In Neu | Ex Neu |
| M01 | 674 | 23ad | $5.2x10^{-59}$ | NS | $9.1x10^{-57}$ | $6.8x10^{-44}$ | NS | NS |
| M02 | 1501 | 33ad | NS | NS | NS | 0.02 | 0.02 | NS |
| M03 | 112 | 33ad | $8.4x10^{-3}$ | $5.5x10^{-6}$ | $1.9x10^{-43}$ | $3.2x10^{-6}$ | NS | NS |
| M04 | 203 | 33ad | $1.1x10^{-6}$ | NS | $2.8x10^{-5}$ | $8.7x10^{-9}$ | NS | NS |
| M05 | 449 | 33ad | NS | $1.6x10^{-7}$ | NS | $5.3x10^{-10}$ | $1.1x10^{-4}$ | $2.8x10^{-18}$ |
| M06 | 495 | 33ad | $2.6x10^{-97}$ | 0.01 | $1.1x10^{-12}$ | $2.4x10^{-44}$ | NS | NS |
| M07 | 774 | 33ad | NS | 0.02 | NS | $1.2x10^{-5}$ | $5.2x10^{-14}$ | $7.6x10^{-147}$ |
| M08 | 247 | 33ad | NS | $8.1x10^{-5}$ | $1.7x10^{-6}$ | NS | NS | NS |
| M09 | 546 | 33ad | NS | NS | NS | $3.6x10^{-6}$ | $1.3x10^{-41}$ | $1.6x10^{-313}$ |
| M10 | 2105 | 34ad | NS | $1.0x10^{-3}$ | NS | $6.1x10^{-14}$ | $3.3x10^{-41}$ | $2.6x10^{-277}$ |
| M11 | 1242 | 34ad | NS | NS | NS | $6.9x10^{-3}$ | NS | NS |
| M12 | 407 | 34ad | $1.0x10^{-22}$ | $4.9x10^{-3}$ | $5.0x10^{-8}$ | $5.6x10^{-20}$ | NS | NS |
| M13 | 491 | 34ad | NS | $1.7x10^{-3}$ | $5.3x10^{-3}$ | $3.5x10^{-12}$ | $1.9x10^{-7}$ | $2.6x10^{-66}$ |
| M14 | 314 | 33c | NS | NS | NS | 0.02 | $3.9x10^{-9}$ | $7.4x10^{-96}$ |
| M15 | 707 | 33c | NS | NS | NS | $5.0x10^{-3}$ | NS | NS |
| M16 | 779 | 33c | NS | NS | NS | $5.7x10^{-7}$ | $4.5x10^{-40}$ | 0 |
| M17 | 198 | 33c | $6.9x10^{-5}$ | $7.4x10^{-11}$ | $1.4x10^{-29}$ | $1.1x10^{-9}$ | 0.03 | NS |
| M18 | 910 | 33c | NS | NS | NS | NS | $4.7x10^{-8}$ | $2.2x10^{-5}$ |
| M19 | 599 | 33c | $3.2x10^{-118}$ | $4.5x10^{-4}$ | $8.0x10^{-16}$ | $3.1x10^{-32}$ | NS | NS |
| M20 | 2234 | 33c | NS | NS | NS | NS | $6.6x10^{-10}$ | $6.8x10^{-64}$ |
| M21 | 320 | 34c | NS | $3.6x10^{-6}$ | NS | $1.7x10^{-8}$ | $2.3x10^{-3}$ | $5.5x10^{-29}$ |
| M22 | 254 | 34c | NS | NS | NS | NS | NS | $9.1x10^{-38}$ |
| M23 | 782 | 34c | NS | NS | NS | $2.9x10^{-6}$ | $3.1x10^{-38}$ | $2.8x10^{-277}$ |

NS: not significant; Ast: astrocytes; Ex Neu: excitatory neurons; In Neu: inhibitory neurons; Mic: microglia; Oli: oligodendrocytes; OPC: oligodendrocyte progenitor cells

**Supplementary Table 10. Association of Expression of Network M01 Genes with Braak Stage and Plaque Score**

| Gene | Braak Stage | | | | | | Plaque Score | | | | | |
|---|---|---|---|---|---|---|---|---|---|---|---|---|
| | ROSMAP | | FHS/BUADC | | Meta-Analysis | | ROSMAP | | FHS/BUADC | | Meta-Analysis | |
| | β | P-value | β | P-value | β | P-value | β | P-value | β | P-value | β | P-value |
| *ARHGEF37* | 0.23 | $6.3x10^{-3}$ | 0.15 | $8.0x10^{-3}$ | 0.17 | $1.7x10^{-4}$ | -0.29 | $7.2x10^{-4}$ | -0.1 | 0.12 | -0.17 | $1.0x10^{-3}$ |
| *C4A* | 0.15 | $5.4x10^{-4}$ | 0.09 | 0.36 | 0.14 | $3.7x10^{-4}$ | -0.13 | $2.8x10^{-3}$ | -0.14 | 0.23 | -0.13 | $1.2x10^{-3}$ |
| *C4B* | 0.16 | $2.6x10^{-4}$ | 0.13 | 0.21 | 0.16 | $1.1x10^{-4}$ | -0.16 | $5.3x10^{-4}$ | -0.19 | 0.11 | -0.16 | $1.3x10^{-4}$ |
| *C5orf64* | 0.08 | 0.17 | 0.1 | 0.15 | 0.09 | 0.05 | -0.11 | 0.09 | -0.07 | 0.31 | -0.09 | 0.05 |
| *CALD1* | 0.18 | 0.06 | 0.09 | 0.02 | 0.1 | $4.2x10^{-3}$ | -0.22 | 0.02 | -0.07 | 0.09 | -0.1 | 0.01 |
| *CHST6* | 0.13 | 0.02 | 0.16 | 0.03 | 0.14 | $1.6x10^{-3}$ | -0.16 | $5.3x10^{-3}$ | -0.16 | 0.05 | -0.16 | $6.2x10^{-4}$ |
| *DOCK1* | 0.17 | 0.05 | 0.06 | 0.12 | 0.08 | 0.02 | -0.23 | $6.6x10^{-3}$ | -0.04 | 0.38 | -0.08 | 0.04 |
| *DOCK5* | 0.19 | 0.01 | 0.1 | 0.15 | 0.14 | $5.9x10^{-3}$ | -0.23 | $2.8x10^{-3}$ | -0.05 | 0.55 | -0.14 | 0.01 |
| *HIP1* | 0.12 | 0.16 | 0.14 | $6.9x10^{-3}$ | 0.13 | $2.1x10^{-3}$ | -0.25 | $3.4x10^{-3}$ | -0.11 | 0.06 | -0.15 | $1.3x10^{-3}$ |
| *HSPA2* | 0.2 | $1.6x10^{-3}$ | 0.14 | 0.09 | 0.18 | $3.9x10^{-4}$ | -0.33 | $4.7x10^{-7}$ | -0.10 | 0.27 | -0.25 | $1.6x10^{-6}$ |
| *LPAR1* | 0.12 | 0.06 | 0.15 | 0.02 | 0.14 | $2.1x10^{-3}$ | -0.23 | $3.7x10^{-4}$ | -0.09 | 0.19 | -0.17 | $4.5x10^{-4}$ |
| *MCM7* | 0.24 | 0.01 | 0.14 | $3.4x10^{-4}$ | 0.15 | $1.4x10^{-5}$ | -0.35 | $3.1x10^{-4}$ | -0.11 | 0.01 | -0.15 | $1.3x10^{-4}$ |
| *PHLPP1* | 0.24 | 0.02 | 0.07 | 0.2 | 0.1 | 0.03 | -0.38 | $3.8x10^{-4}$ | -0.04 | 0.5 | -0.11 | 0.02 |
| *PLEKHA7* | 0.09 | 0.27 | 0.13 | 0.03 | 0.12 | 0.01 | -0.18 | 0.03 | -0.09 | 0.19 | -0.12 | 0.02 |
| *PRIMA1* | 0.15 | 0.03 | 0.13 | 0.15 | 0.14 | $8.8x10^{-3}$ | -0.22 | $1.5x10^{-3}$ | -0.09 | 0.35 | -0.18 | $1.6x10^{-3}$ |
| *RHOU* | 0.21 | $9.2x10^{-3}$ | 0.04 | 0.49 | 0.1 | 0.04 | -0.22 | $6.8x10^{-3}$ | -0.04 | 0.56 | -0.11 | 0.03 |
| *RNF43* | -0.23 | $3.1x10^{-3}$ | -0.07 | 0.3 | -0.13 | $6.6x10^{-3}$ | 0.19 | 0.02 | 0.06 | 0.42 | 0.11 | 0.03 |
| *SLC45A3* | 0.17 | 0.01 | 0.13 | 0.16 | 0.16 | $4.0x10^{-3}$ | -0.29 | $1.9x10^{-5}$ | -0.08 | 0.44 | -0.23 | $4.7x10^{-5}$ |
| *TNS3* | 0.14 | 0.04 | 0.09 | 0.1 | 0.11 | $8.7x10^{-3}$ | -0.18 | $6.2x10^{-3}$ | -0.04 | 0.46 | -0.11 | 0.02 |

Expression of genes listed were nominally associated ($P<0.05$) in both traits in the meta-analysis. Expression differences (β estimate represent the magnitude of effect) for each trait were calculated using a linear regression model separately in each dataset and the results were combined by meta-analysis.

**Supplementary Table 11.** Differential expression of genes from the M01 network between AD cases and controls by brain cell type *

| Gene | Mean Expression | | | Ast | | Oli | | OPC | |
|---|---|---|---|---|---|---|---|---|---|
| | Ast | Oli | OPC | LogFC[1] | P-value | LogFC | P-value | LogFC | P-value |
| *CALD1* | 3.60 | 9.72 | 10.37 | 0.015 | 2.7E-01 | -0.039 | 6.8E-01 | -0.185 | 7.8E-03 |
| *DOCK1* | NA | 2.07 | NA | NA | NA | 0.245 | **3.8E-07** | NA | NA |
| *DOCK5* | NA | 10.99 | 5.07 | NA | NA | 0.075 | 1.8E-05 | 0.039 | 8.0E-01 |
| *HIP1* | NA | 5.06 | 9.13 | NA | NA | 0.026 | 9.3E-03 | -0.013 | 9.0E-01 |
| *HSPA2* | 2.34 | 11.78 | NA | -0.041 | 3.9E-01 | 0.139 | **1.8E-10** | NA | NA |
| *LPAR1* | NA | 4.55 | NA | NA | NA | 0.134 | **7.0E-08** | NA | NA |
| *PHLPP1* | 2.48 | 5.59 | 3.19 | 0.054 | 8.1E-01 | 0.185 | **3.8E-11** | 0.170 | 4.5E-01 |
| *RHOU* | NA | 7.04 | 2.90 | NA | NA | 0.096 | 3.9E-05 | 0.125 | 3.3E-01 |
| *SLC45A3* | NA | 2.14 | NA | NA | NA | 0.041 | 1.5E-02 | NA | NA |
| *TNS3* | 2.59 | NA | NA | 0.014 | 8.0E-01 | NA | NA | NA | NA |

* Genes with low expression (mean < 2) in oligodendrocytes, oligodendrocyte progenitor cells, and astrocytes were excluded.

[1] Log2 fold change of expression levels (LogFC) and P value (P) were calculated between AD and control brains from single nuclei RNA sequencing data from ROSMAP.

Ast: astrocyte; Oli: oligodendrocytes; OPC: oligodendrocyte progenitor cells; NA: not available due to small number of single cells showing expression (average expression < 2).

P values significant after false discovery rate (FDR) adjustment are highlighted in bold.

**Supplementary Table 12. Characteristics of Immunoassay Measures in FHS/BUADC**

| Group | Aβ42 | | pTau181/tTau | | pTau231/tTau | | C4A | | C4B | |
|---|---|---|---|---|---|---|---|---|---|---|
| | N | Mean | N | Mean | N | Mean | N | Mean | N | Mean |
| Total | 191 | $8.7 \times 10^{-3}$ | 146 | $-7.5 \times 10^{-3}$ | 186 | 0.04 | 179 | $-4.3 \times 10^{-3}$ | 179 | -0.02 |
| ε2/ε3 | 37 | -0.47 | 29 | -0.02 | 37 | -0.23 | 34 | -0.21 | 34 | -0.07 |
| ε3/ε3 | 109 | 0.03 | 81 | 0.02 | 104 | 0.05 | 101 | $7.6 \times 10^{-3}$ | 101 | 0.03 |
| ε3/ε4 | 31 | 0.38 | 25 | -0.05 | 31 | 0.30 | 30 | 0.23 | 30 | -0.16 |

N: sample size; SD: standard deviation; $A\beta_{42}$: amyloid-β 42; pTau181/tTau: ratio of phosphorylated tau at position 181 to total tau; pTau231/tTau: ratio of phosphorylated tau at position 231 to total tau; C4A: complement 4A protein; C4B: complement 4B protein; PSD95: postsynaptic density of 95k Dalton. All traits were rank-transformed for age of death and sex.

**Supplementary Table 13. Association of Expression of *APOE* ε2/ε3-related Genes with Measures of AD-related Proteins ***

| Gene | Aβ42 | | pTau181/tTau | | pTau231/tTau | | PSD95 | | C4A | | C4B | |
|---|---|---|---|---|---|---|---|---|---|---|---|---|
| | β | P | β | P | β | P | β | P | β | P | β | P |
| *C4A* | 0.08 | 0.18 | $-9.1x10^{-3}$ | 0.89 | 0.22 | $5.1x10^{-5}$ | -0.10 | 0.08 | 0.06 | 0.26 | 0.06 | 0.33 |
| *C4B* | 0.08 | 0.17 | $-7.4x10^{-3}$ | 0.91 | 0.21 | $7.0x10^{-5}$ | -0.10 | 0.08 | 0.04 | 0.44 | 0.05 | 0.43 |
| *GFAP* | 0.05 | 0.55 | $-5.6x10^{-3}$ | 0.95 | 0.23 | $1.5x10^{-3}$ | -0.12 | 0.13 | -0.05 | 0.54 | 0.08 | 0.31 |
| *NPNT* | 0.25 | 0.02 | 0.07 | 0.55 | 0.35 | $4.9x10^{-4}$ | -0.08 | 0.48 | 0.15 | 0.16 | -0.14 | 0.23 |
| *DOCK1* | 0.07 | 0.64 | $-1.4x10^{-3}$ | 0.99 | 0.28 | 0.04 | -0.20 | 0.18 | -0.10 | 0.46 | 0.39 | $8.3x10^{-3}$ |
| *HSPA2* | 0.03 | 0.70 | -0.01 | 0.86 | 0.30 | $4.3x10^{-6}$ | -0.10 | 0.17 | 0.10 | 0.13 | 0.20 | $6.1x10^{-3}$ |
| *LPAR1* | -0.07 | 0.49 | -0.02 | 0.88 | -0.05 | 0.60 | $-6.1x10^{-3}$ | 0.95 | -0.08 | 0.39 | 0.18 | 0.07 |
| *PHLPP1* | 0.08 | 0.48 | -0.10 | 0.44 | 0.38 | $3.9x10^{-4}$ | -0.18 | 0.13 | 0.16 | 0.15 | 0.21 | 0.08 |

* Includes significantly differentially expressed genes shown in Table S 1 and Supplemental Table S 11.

Aβ42: amyloid-$\beta_{42}$; pTau181/tTau: ratio of phosphorylated tau at position 181 to total tau; pTau231/tTau: ratio of phosphorylated tau at position 231 to total tau; C4A: complement 4A protein; C4B: complement 4B protein; PSD95: postsynaptic density of 95k Dalton.

Effect size (β) of the association gene expression with protein measures were calculated using a linear

**Supplementary Figure 1.** Quality control of FHS/BUADC RNA sequencing data including the distribution among samples of (A) library size and (B) expression counts across batches. (C) First 2 principle components of gene expression for samples colored by (C) library batches and (D) sequencing batches.

**A.**

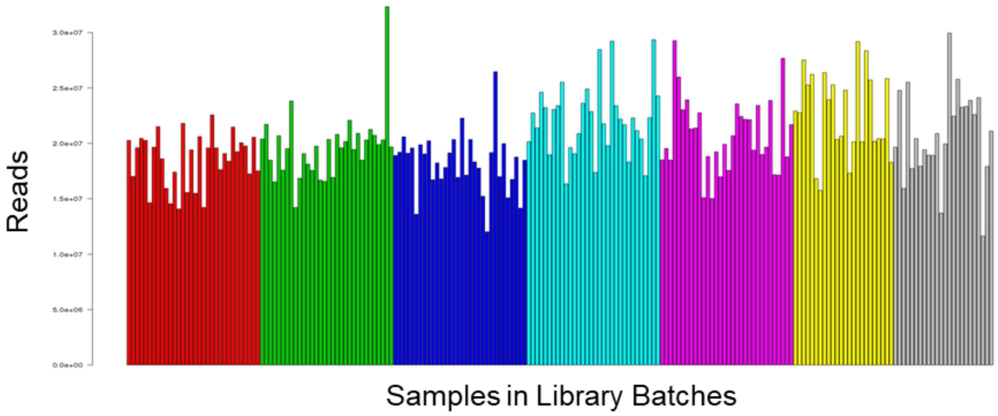


**B.**

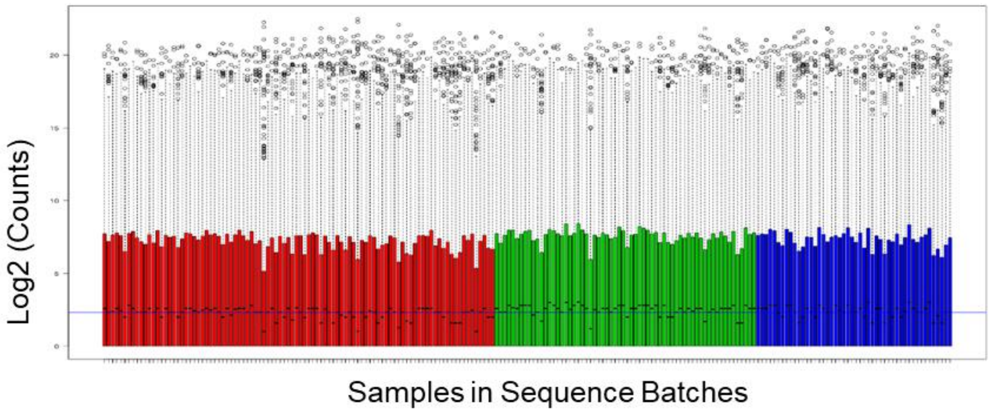


**C.**

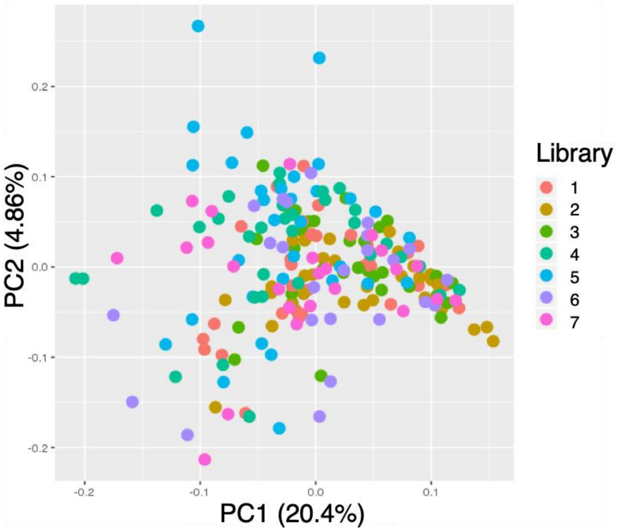


**D.**

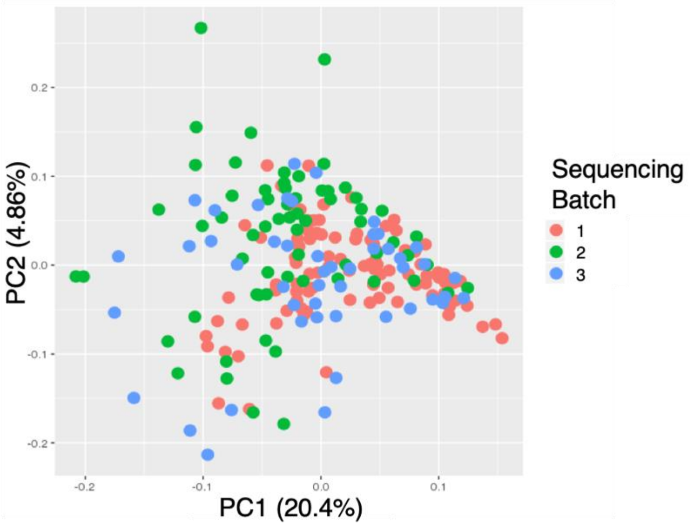

**Supplementary Figure 2. Volcano Plots for Differential Gene Expression Analysis in the ROSMAP Sample Among Subjects with *APOE* Genotypes ε2/ε3 (panel A), ε3/ε3 (panel B) and ε3/ε4 (panel C), and in the Total Sample (panel D).** Measure of differential expression between AD cases and controls expressed as $Log_2$ fold change is shown on the x-axis and significance level expressed as the -log P-value is shown on the y-axis. Results for each gene are represented by colored dots and are restricted to those with | $log_2$ fold-change | > 0.3 or p-value < 0.01. Genes that were surpassed the p-value and $log_2$ fold-change thresholds are shown in red, genes that surpassed the p-value threshold only are shown in blue, genes that surpassed the $log_2$ fold-change threshold only are shown in green, and genes that did not surpass either threshold are shown in grey.

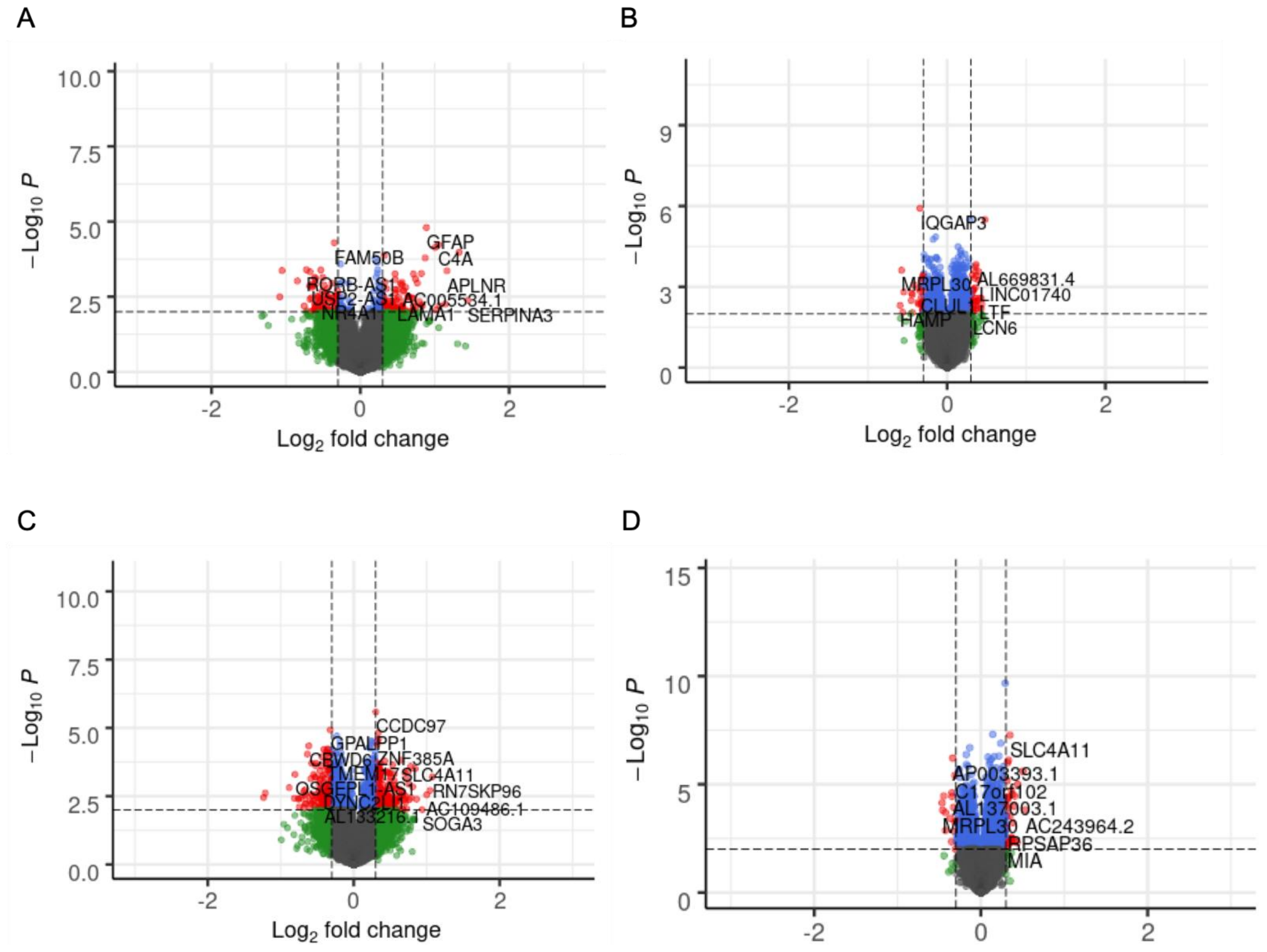

**Supplementary Figure 3. Volcano Plots for Differential Gene Expression Analysis in the MAYO Sample Among Subjects with *APOE* Genotypes ε2/ε3 (panel A), ε3/ε3 (panel B) and ε3/ε4 (panel C), and in the Total Sample (panel D).** Measure of differential expression between AD cases and controls expressed as $Log_2$ fold change is shown on the x-axis and significance level expressed as the -log P-value is shown on the y-axis. Results for each gene are represented by colored dots and are restricted to those with | $log_2$ fold-change | > 0.3 or p-value < 0.01. Genes that were surpassed the p-value and $log_2$ fold-change thresholds are shown in red, genes that surpassed the p-value threshold only are shown in blue, genes that surpassed the $log_2$ fold-change threshold only are shown in green, and genes that did not surpass either threshold are shown in grey.

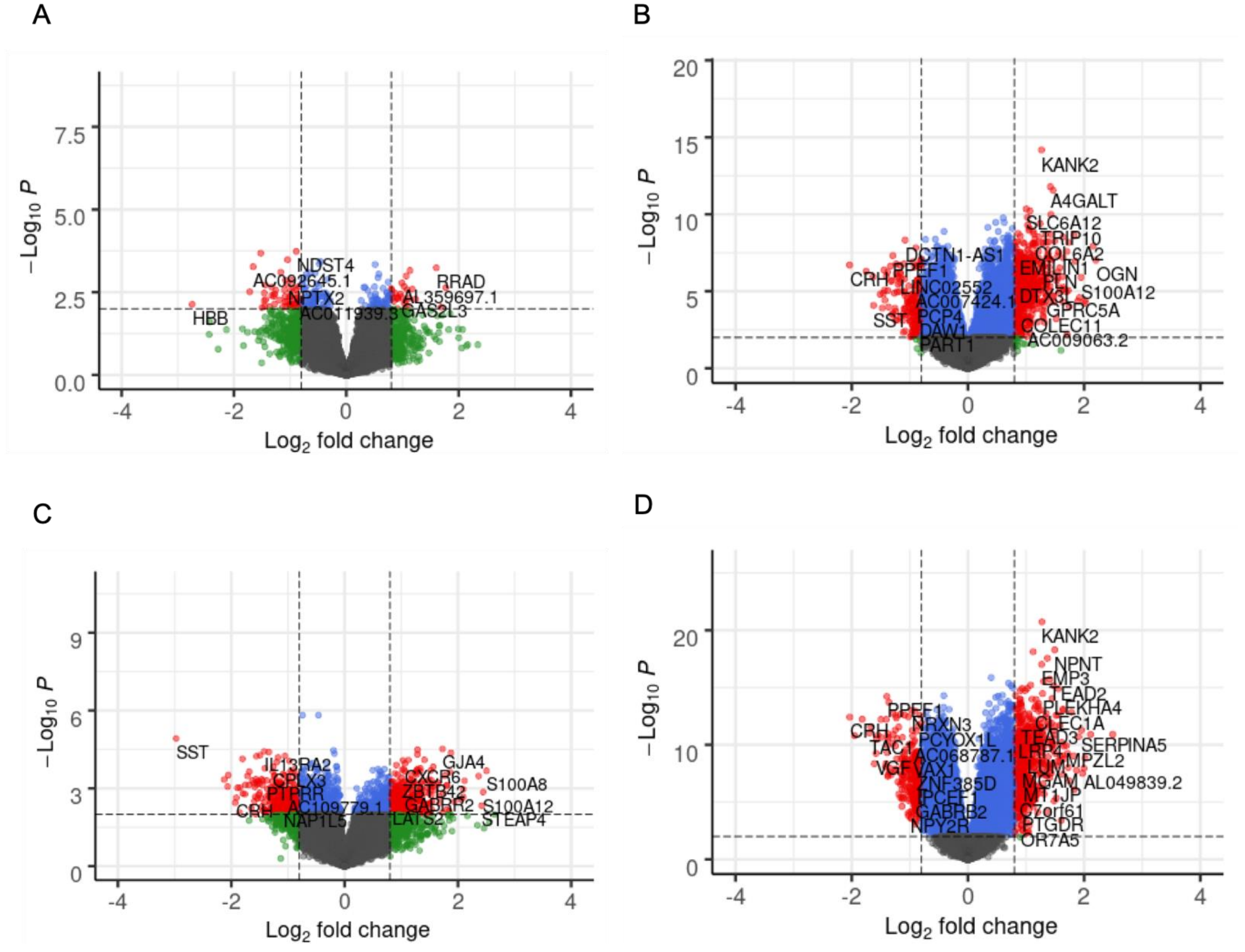

**Supplementary Figure 4. Volcano Plots for Differential Gene Expression Analysis in the FHS/BUADC Sample Among Subjects with *APOE* Genotypes ε2/ε3 (panel A), ε3/ε3 (panel B) and ε3/ε4 (panel C), and in the Total Sample (panel D).** Measure of differential expression between AD cases and controls expressed as $Log_2$ fold change is shown on the x-axis and significance level expressed as the -log P-value is shown on the y-axis. Results for each gene are represented by colored dots and are restricted to those with | $log_2$ fold-change | > 0.3 or p-value < 0.01. Genes that were surpassed the p-value and $log_2$ fold-change thresholds are shown in red, genes that surpassed the p-value threshold only are shown in blue, genes that surpassed the $log_2$ fold-change threshold only are shown in green, and genes that did not surpass either threshold are shown in grey.

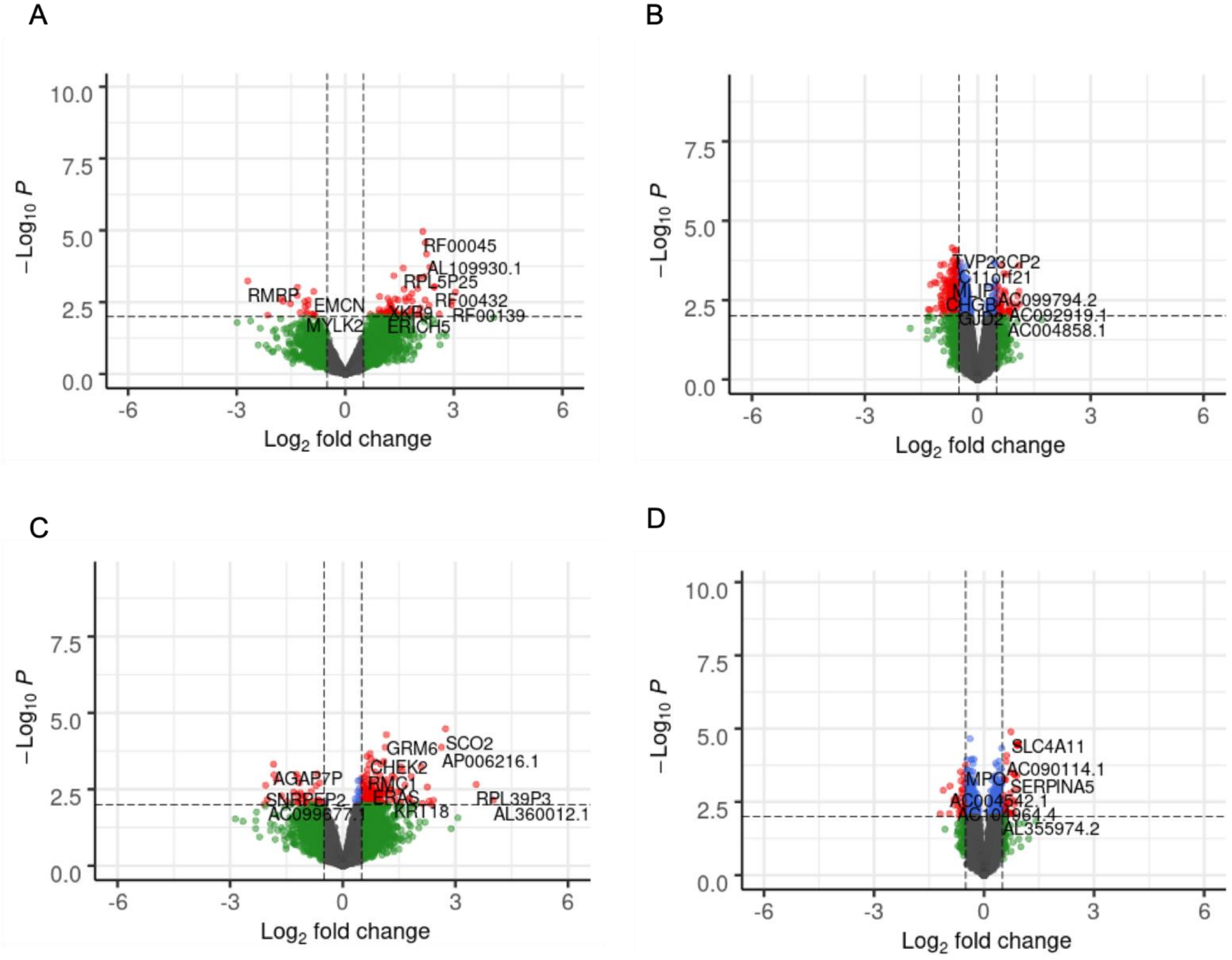

**Supplementary Figure 5.** Number of significant differentially expressed genes (p<0.01) in each of and across the *APOE* genotype groups (ε2/ε3, ε3/ε3, ε3/ε4) and the total sample.

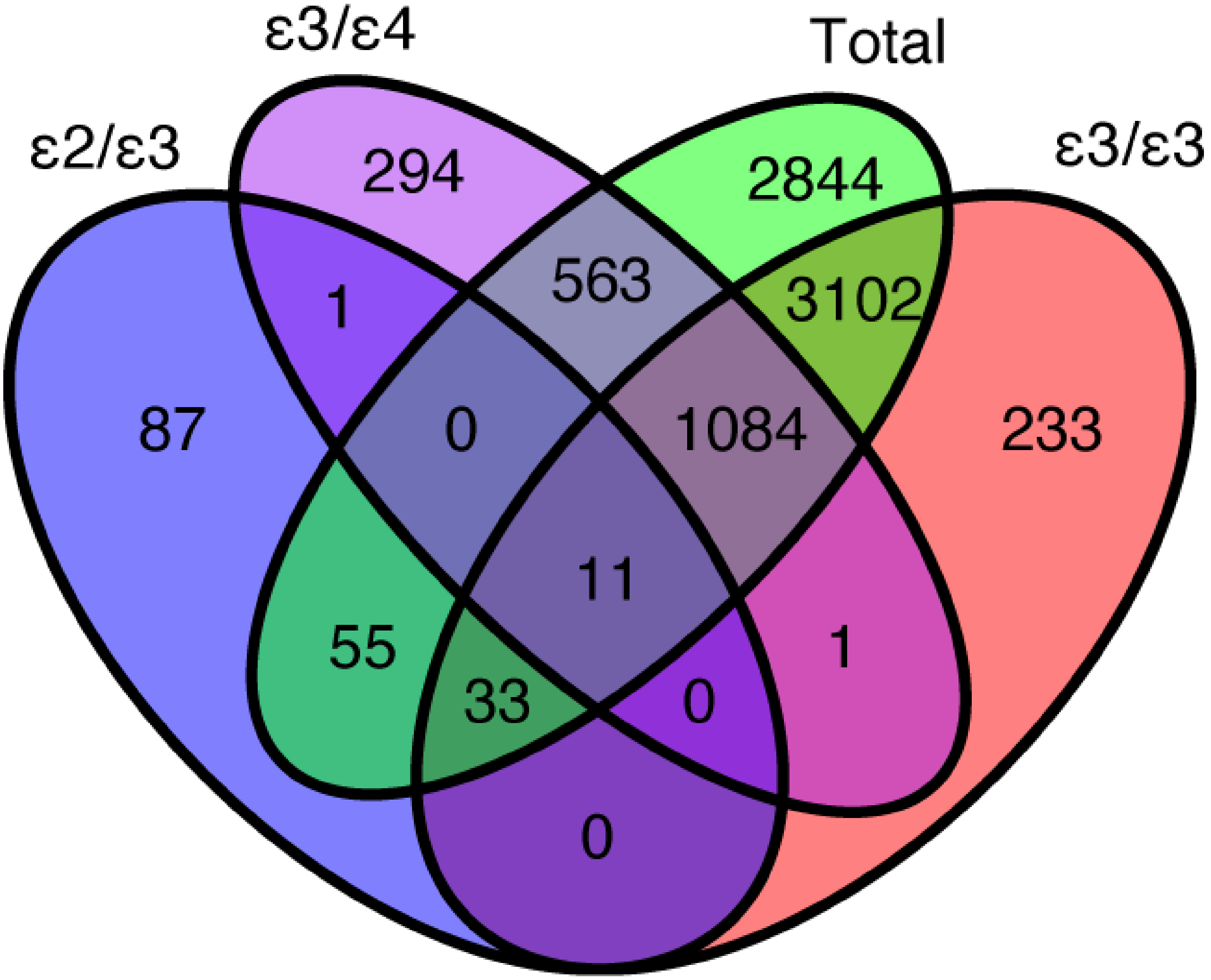

**Supplementary Figure 6.** Cell-type characterization of brain tissue obtained from 24 AD cases and 24 controls in the ROSMAP Study. Plot shows a two-dimensional t-distributed stochastic neighbor embedding (t-SNE) projection of all annotated cells derived from single nuclei RNA sequencing (snRNA-seq) data.

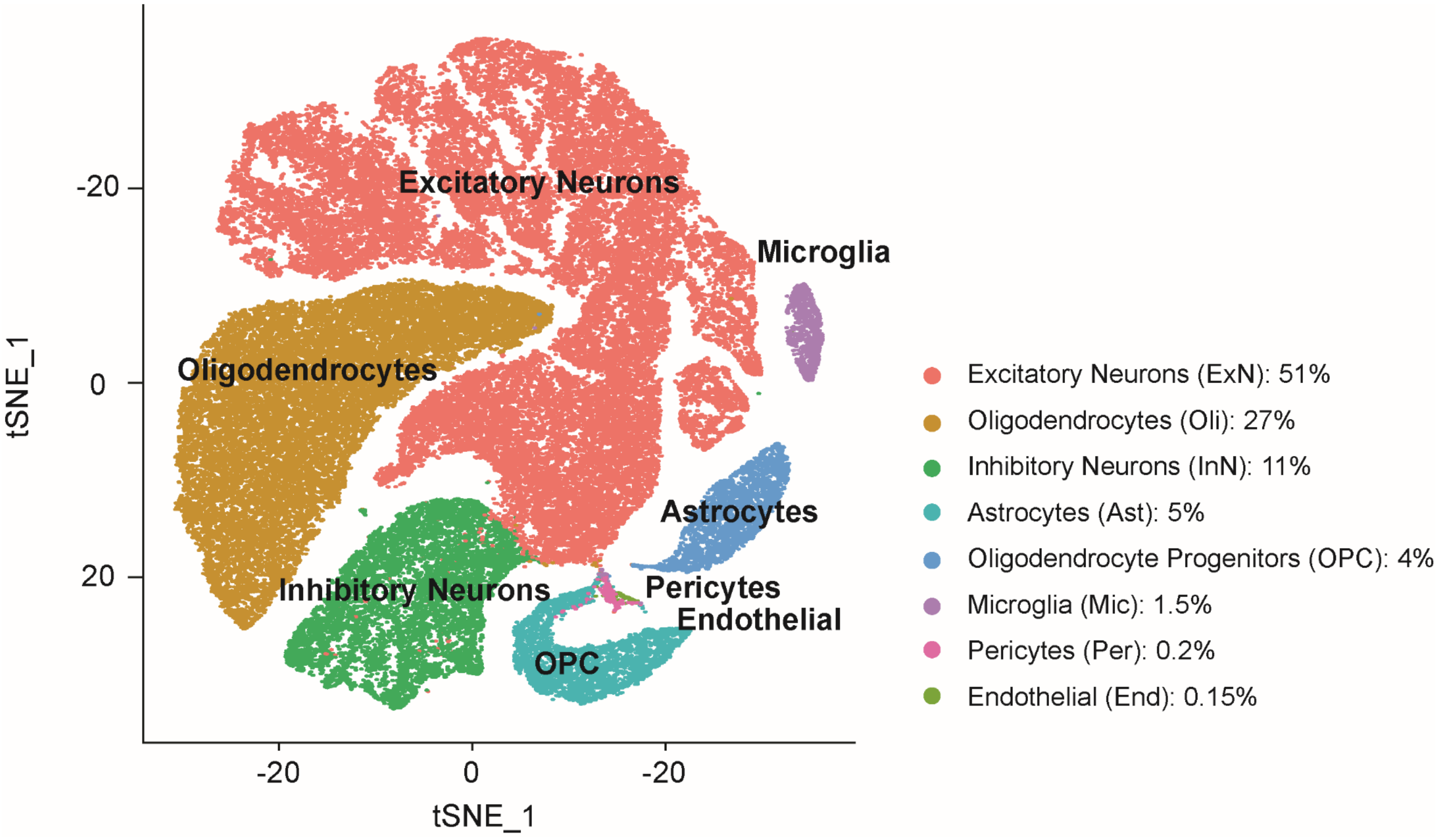

**Supplementary Figure 7.** Cell type-specific marker genes. (A) *SYT1* for neurons, (B) *GAD1* for excitatory neurons, (C) *SLC17A7* for inhibitory neurons, (D) *PLP1* for oligodendrocytes, (E) *AQP4* for oligodendrocyte progenitor cells, (F) *VCAN* for astrocytes, and (G) *CD74* for microglia.

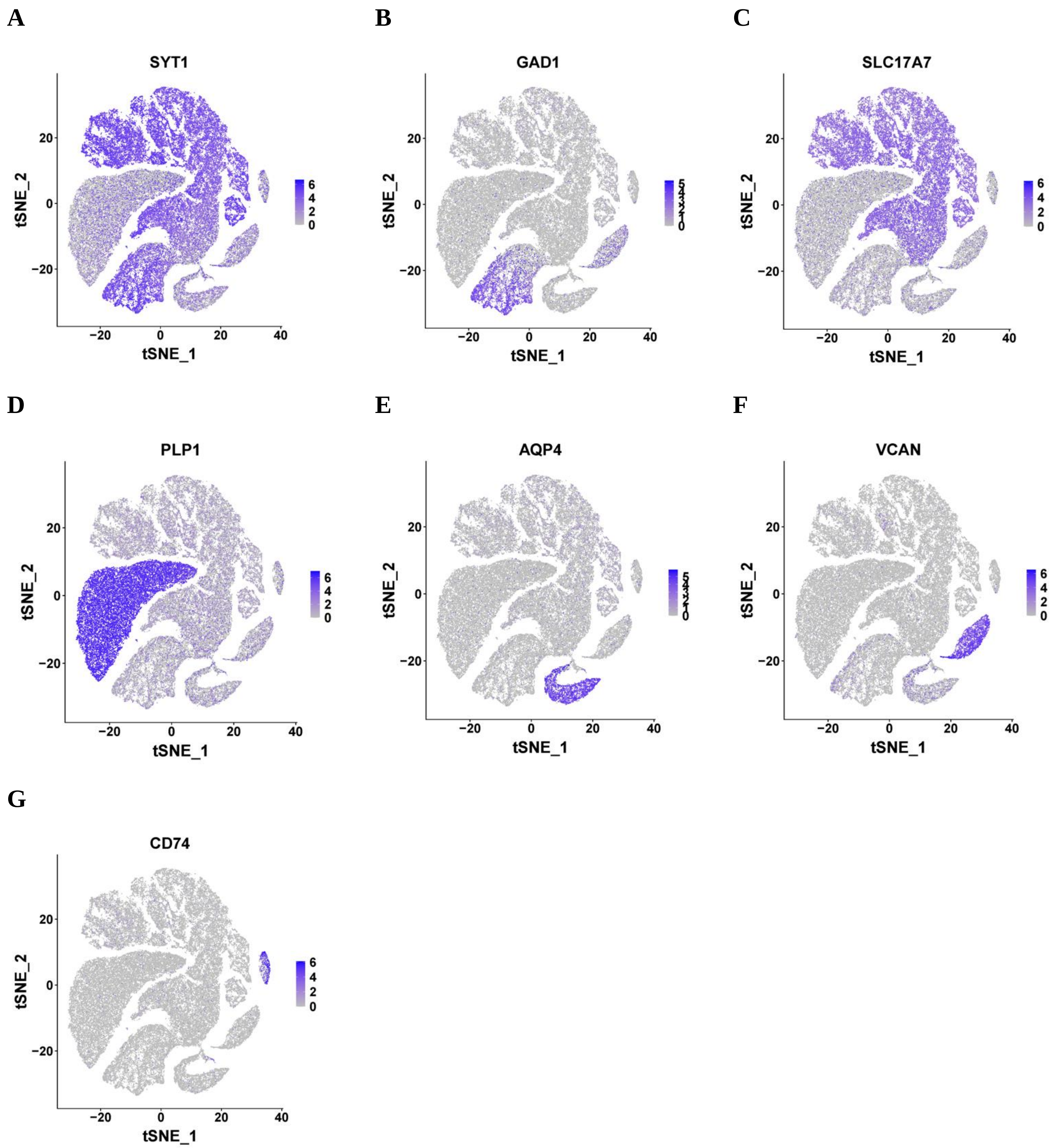

**Supplementary Figure 8.** Cell-type distribution of co-expressed genes in *APOE* ε2/ε3 AD cases reported in Supplementary Table 13.

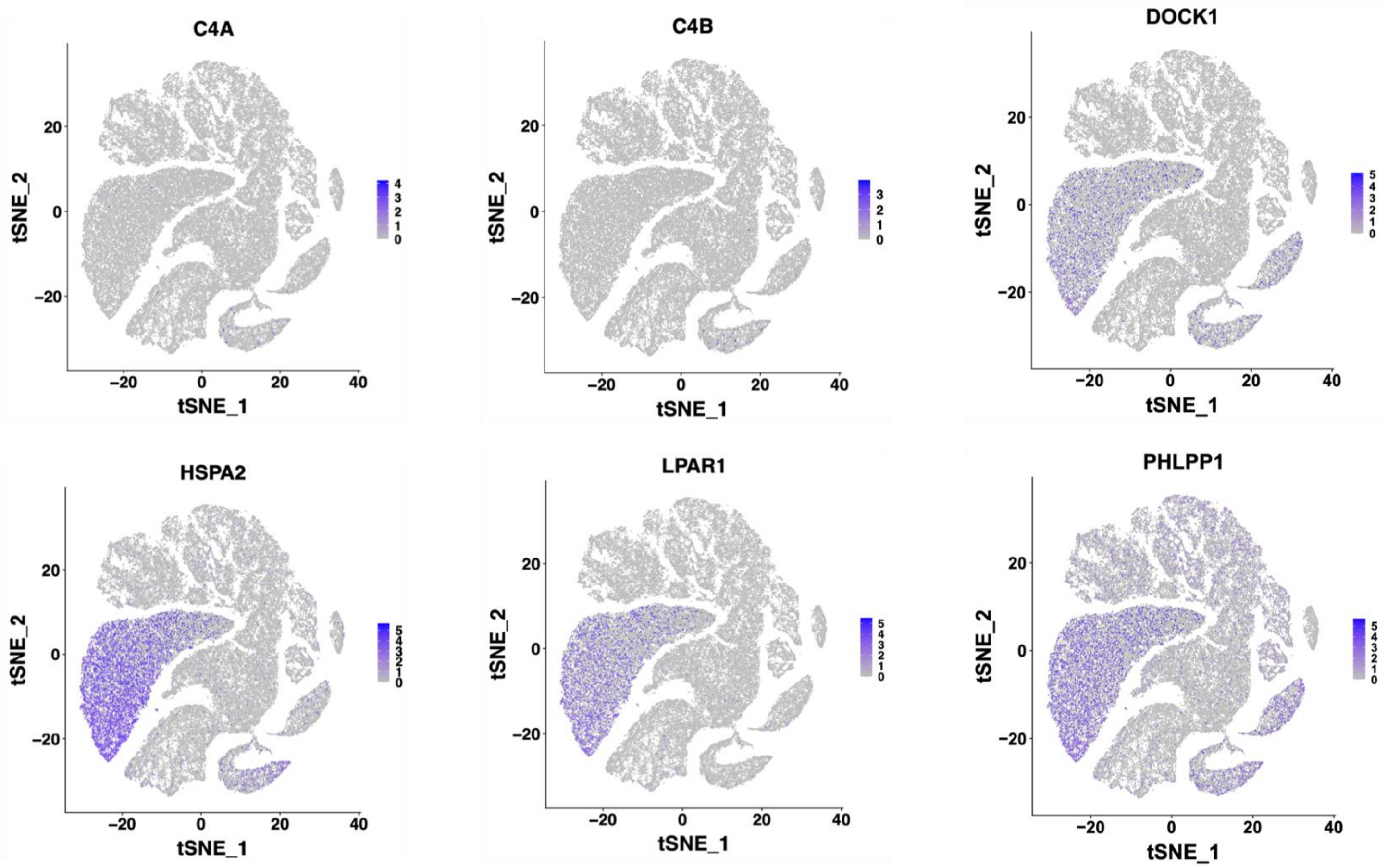
